# Clinically Informed Semi-Supervised Learning Improves Disease Annotation and Equity from Electronic Health Records: A Glaucoma Case Study

**DOI:** 10.1101/2025.09.12.25335665

**Authors:** Mousa Moradi, Rishi Shah, Asahi Fujita, Niloufar Bineshfar, Daniel M. Vu, Kanza Aziz, Daniel L. Liebman, Saber Kazeminasab Hashemabad, Mengyu Wang, Tobias Elze, Mohammad Eslami, Nazlee Zebardast

## Abstract

Clinical notes represent a vast but underutilized source of information for disease characterization, whereas structured electronic health record (EHR) data such as ICD codes are often noisy, incomplete, and too coarse to capture clinical complexity. These limitations constrain the accuracy of datasets used to investigate disease pathogenesis and progression and to develop robust artificial intelligence (AI) systems. To address this challenge, we introduce Ci-SSGAN (Clinically Informed Semi-Supervised Generative Adversarial Network), a novel framework that leverages large-scale unlabeled clinical text to reannotate patient conditions with improved accuracy and equity. As a case study, we applied Ci-SSGAN to glaucoma, a leading cause of irreversible blindness characterized by pronounced racial and ethnic disparities. Trained on 2.1 million ophthalmology notes, Ci-SSGAN achieved 0.85 accuracy and 0.95 AUROC, representing a 10.19% AUROC improvement compared to ICD-based labels (0.74 accuracy, 0.85 AUROC). Ci-SSGAN also narrowed subgroup performance gaps, with F1 gains for Black patients (+0.05), women (+0.06), and younger patients (+0.033). By integrating semi-supervised learning and demographic conditioning, Ci-SSGAN minimizes reliance on expert annotations, making AI development more accessible to resource-constrained healthcare systems.

## Introduction

Artificial intelligence (AI) and large language models (LLMs) are transforming healthcare by uncovering insights from complex medical data that are often inaccessible in routine clinical decision-making. Domain-specific language models such as BioClinical BERT ^1-3^, Med-PaLM ^4,5^, MedGemma ^6^, and GPT ^7-10^ have demonstrated remarkable capabilities in processing clinical text, enabling applications from disease classification to clinical reasoning and question-answering.

Approximately 80% of clinically relevant information in electronic health records (EHRs) is found in unstructured clinical notes, which capture nuanced patient presentations, physician reasoning, and disease progression in ways that structured labels, such as ICD codes, often fail to represent ^11^. Despite their richness, clinical notes remain largely underutilized for large-scale research and model development ^12^. In contrast, ICD codes and other structured EHR labels are widely used but are known to be noisy ^13^, incomplete, and often too coarse for tasks requiring fine-grained annotation. This mismatch leaves researchers with inaccurate datasets that limit both clinical insights and the reliability of AI systems built upon them.

Clinical practice generates vast amounts of unlabeled notes, and our tertiary academic center institution alone has over 250 million, including two million ophthalmology records (as illustrated in Fig. 1a). Obtaining expert labels at this scale is impractical, leaving abundant data largely unusable for supervised learning. Expert clinical grading remains the gold standard for disease classification, yet obtaining such labels at scale is costly and time-consuming. However, fully supervised models, including BERT variants ^1,2^, require large, high-quality labeled datasets that are rarely available for specialized disease classification tasks.

**Fig. 1.**
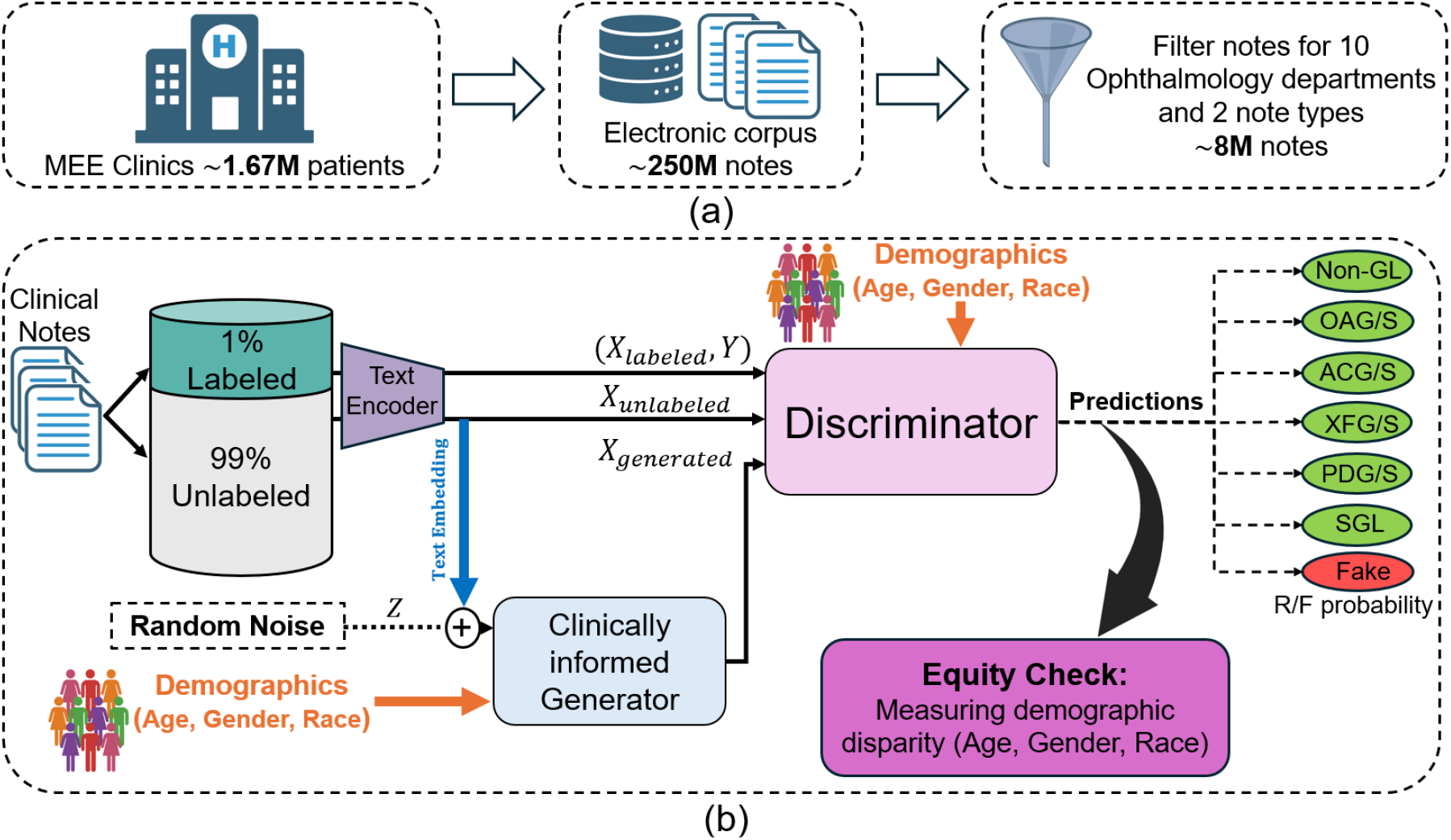
Overview of the Ci-SSGAN framework. **a** Abundant unlabeled ophthalmology notes compared with limited expert annotations. **b** Ci-SSGAN generator combines unlabeled text embeddings (blue arrow), demographics (orange arrow), and noise (dashed black arrow) to produce synthetic data, with the discriminator classifying glaucoma subtypes and output evaluated to ensure fairness by measuring each subgroup disparities. Non-GL=non-glaucoma, OAG/S= open angle glaucoma/suspect, ACG/S= angle closure glaucoma/suspect, XFG/S= exfoliation glaucoma/syndrome, PDG/S= pigmentary dispersion glaucoma/syndrome, and SGL= secondary glaucoma. R= real, F= fake.

Furthermore, EHR labels in their current form can reinforce biases and overlook important subgroups, especially for diseases with complex subtypes or variable presentations. When combined with limited labeled data, these biases reduce model reliability and underscore the need for frameworks that harness unstructured clinical data while promoting equitable performance. When training data lacks demographic diversity or contains label noise, supervised models perform substantially worse ^11,14,15^. For example, prior studies have shown that feature-dependent label noise, annotator disagreements, or limited diversity in training corpora can degrade performance, and these effects are not fully corrected by fine-tuning ^16,17^.

To address label scarcity, systemic biases, and the underutilization of abundant unlabeled clinical notes, we propose Ci-SSGAN (Clinically Informed Semi-Supervised Generative Adversarial Network), a framework explicitly designed to ensure equitable performance across demographic groups (as illustrated in Fig. 1b). The framework achieves higher accuracy with substantially fewer labeled examples. It ensures reliable outcomes and reduces disparities by conditioning semi-supervised generation on unlabeled clinical text and patient characteristics. Unlike standard semi-supervised GANs ^18,19^ which use only noise as their generator inputs, Ci-SSGAN introduces three key innovations: (1) a clinically informed generator that integrates embeddings from unlabeled clinical text with demographic conditions and noise, creating synthetic data that is both clinically meaningful and demographically representative; (2) multi-conditional learning that explicitly incorporates race, gender, and age to combat dataset imbalance and ensure consistent performance across all subgroups; and (3) a systematic equity framework using our proposed Parity Violation (PV) score to quantify and minimize disparities in positive and negative predictive values across demographics.

We evaluated Ci-SSGAN in glaucoma subtype detection, a disease that highlights both the opportunities and challenges of AI in healthcare. Glaucoma is the leading cause of irreversible blindness worldwide, projected to affect 80 million people by 2040 with considerable racial, ethnic and socioeconomic disparities making it a compelling test case for equitable AI. Additionally, ICD coding for glaucoma classification has particularly poor specificity ^20^ (less than 50%) and achieves only ∼81% accuracy ^21^, with systematic undercoding, overcoding, and subtype misclassification that undermine reliable labeling. Glaucoma exemplifies both the global burden of disease and the equity challenges that Ci-SSGAN aims to address. Pronounced disparities exist across race, gender, and age groups ^22-29^, making it a compelling real-world test case for equitable AI.

This combination of global burden, pronounced disparities and imperfect structured labels, makes glaucoma an ideal real-world setting to evaluate Ci-SSGAN. By leveraging over two million unlabeled ophthalmology notes alongside a smaller expert-annotated set, our framework demonstrates how semi-supervised learning can address the labeled-data bottleneck while promoting equitable performance across subgroups. Although glaucoma serves as our case study, the approach can be generalized to other diseases where unlabeled notes are abundant but equitable performance remains elusive.

## Results

### Patients Demographics and Clinical Characteristics

The labeled dataset comprised 2954 notes for 1105 patients (53.5% female) and the unlabeled dataset contained 349587 notes for 108574 patients (59% female) who received care at the Massachusetts Eye and Ear (MEE) ophthalmology department from May 2015 to December 2024.

The participants’ age at the time of study ranged from 30 to 90 years for both datasets, with median of 62 and 68 for labeled and unlabeled datasets, respectively. The patients’ demographics in the labeled dataset include 41.7% White or Caucasian, 29.6% Black or African American, and 28.7% Asian. The unlabeled dataset showed similar racial distribution with 40.2% White or Caucasian, 19.5% Black or African American, and 40.3% Asian patients. Table 1 shows demographics and clinical characteristics of the patients in both datasets.

**Table 1.** Demographics and baseline characteristics of the patients are included in the study. The data are presented in form of n(%) or mean ± SD.

| Characteristics | Labeled Dataset | Unlabeled Dataset |
| --- | --- | --- |
| Number of patients | 1105 | 108574 |
| Number of notes | 2954 | 349587 |
| Age (years) |  |  |
| Mean ± SD | 60.4 ± 15.5 | 65.4 ± 15.0 |

| Median (range) | 62 (30-90) | 68 (30-90) |
| --- | --- | --- |
| Age groups, n (%) |  |  |
| 30-55 | 131 (11.9) | 21134 (19.5) |
| 55-70 | 374 (33.8) | 40436 (37.2) |
| >=70 | 600 (54.3) | 47004 (43.3) |
| Gender, n (%) |  |  |
| Female | 591 (53.5) | 64059 (59) |
| Race, n (%) |  |  |
| Asian | 317 (28.7) | 43764 (40.3) |
| Black or African American | 327 (29.6) | 21174 (19.5) |
| White or Caucasian | 461 (41.7) | 43636 (40.2) |

N= number of patients.

### Model performance and equity analysis

To comprehensively demonstrate the advantages of our proposed semi-supervised learning method, We trained all methods with 25%, 50%, and 100% of the labeled notes and evaluated at the patient level. Unless stated otherwise, higher is better for Accuracy/F1/AUROC/AUPRC, and lower is better for the Parity-Violation (PV) score. For model training, the labeled dataset was split into 90% for training and validation (1000 patients, 2660 notes) and 10% used as a held-out test set (105 patients, 294 notes). We evaluated the performance of Ci-SSGAN at each labeled data fraction against two fully supervised models (Base BERT ^1^ and BioClinical BERT ^2^), as well as the standard SSGAN ^18,19^. For classification, we defined six outcome classes: non-glaucoma (Non-GL), open-angle glaucoma/suspect (OAG/S), angle-closure glaucoma/suspect (ACG/S), exfoliation glaucoma/syndrome (XFG/S), pigmentary glaucoma/syndrome (PDG/S), and secondary glaucoma (SGL). These categories were chosen to reflect clinically relevant subtypes with sufficient representation for model training and evaluation.

Performance improvements with Ci-SSGAN were consistent in both note-level and patient-level analyses (see Fig. 2, Table 2 and Supplementary Figures 1, 2), confirming the robustness of its gains over Base BERT, Bio BERT, and standard SSGAN regardless of evaluation granularity. Across all labeled data fractions, Ci-SSGAN achieved the highest Accuracy, AUC-PR, and AUCROC, with improvements over Base BERT and Bio BERT particularly pronounced in low-data settings (AUC-PR +0.091 and +0.092 at 25%). Standard deviations across 5 folds were consistently low, indicating stable performance across runs.

**Table 2.** Class-wise comparison of all trained models on different data fractions. Values are presented in form of Mean± SD. Bold values show the best scores. Standard deviations are calculated across 5 folds.

| Labeled data Fraction | Model | Accuracy | AUC-PR | AUCROC | PV score |
| --- | --- | --- | --- | --- | --- |
| 25%<br>(250 patients with 3081 total notes) | Ci-SSGAN | **\* 0.871 ± 0.03** | **0.893 ± 0.03** | 0.956 ± 0.01 | **0.029** |
|  | SSGAN | 0.859 ± 0.04 | 0.802 ± 0.06 | 0.941 ± 0.03 | 0.031 |
|  | Base BERT | 0.824 ± 0.05 | 0.867 ± 0.05 | 0.957 ± 0.04 | 0.127 |
|  | Bio BERT | 0.781 ± 0.06 | 0.801 ± 0.04 | 0.907 ± 0.03 | 0.104 |
| 50%<br>(500 patients with 6162 notes) | Ci-SSGAN | **0.883 ± 0.02** | **0.901 ± 0.01** | **0.963 ± 0.02** | **0.024** |
|  | SSGAN | 0.849 ± 0.03 | 0.851 ± 0.05 | 0.957 ± 0.04 | 0.034 |
|  | Base BERT | 0.851 ± 0.04 | 0.870 ± 0.02 | 0.954 ± 0.03 | 0.081 |
|  | Bio BERT | 0.801 ± 0.01 | 0.845 ± 0.06 | 0.941 ± 0.04 | 0.075 |
| 100%<br>(1000 patients with 12324 notes) | Ci-SSGAN | **0.897 ± 0.02** | **0.904 ± 0.03** | 0.968 ± 0.01 | **0.022** |
|  | SSGAN | 0.847 ± 0.05 | 0.885 ± 0.06 | 0.967 ± 0.03 | 0.037 |
|  | Base BERT | 0.871 ± 0.07 | 0.882 ± 0.04 | 0.953 ± 0.03 | 0.029 |
|  | Bio BERT | 0.826 ± 0.06 | 0.888 ± 0.02 | 0.969 ± 0.05 | 0.053 |

\*Mean± SD

**Fig. 2.**
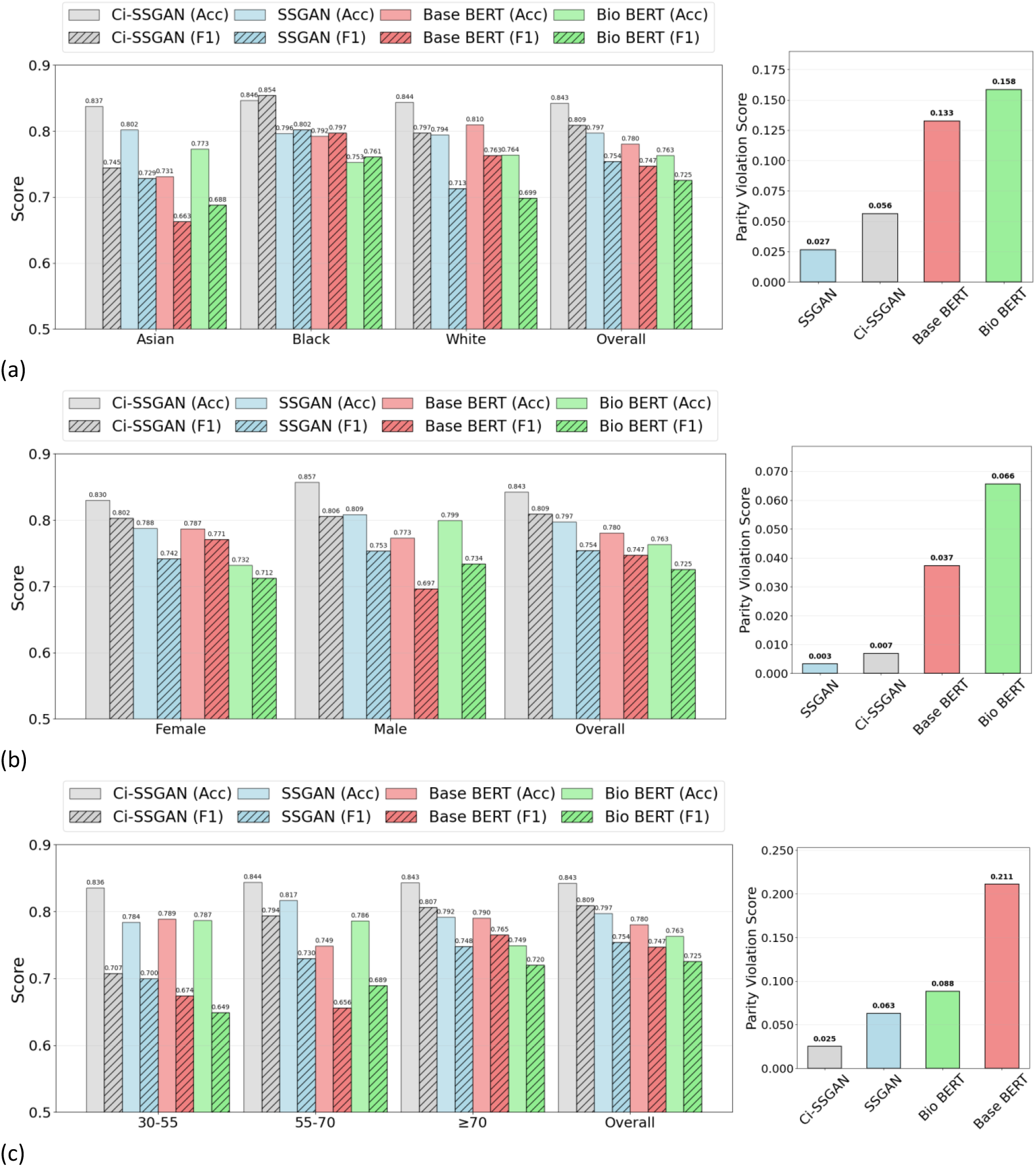
Performance comparison of Ci-SSGAN, SSGAN, Base BERT, and Bio BERT models across: **a** Racial groups, **b** Gender groups, and **c** Age groups using 25% of the labeled data. Left panels show accuracy (solid bars) and F1-score (hatched bars) for each subgroup and overall performance. Right panels present corresponding parity violation scores, indicating fairness across demographic subgroups. Ci-SSGAN consistently achieves higher accuracy and F1-scores with lower parity violations compared to other models. The results are presented on five CV folds. Acc= Accuracy, F1= F1-macro.

When utilizing only 25% of labeled data (Fig. 2), Ci-SSGAN achieved notable gains over Base BERT, increasing overall accuracy from 0.78 to 0.843 and F1 score from 0.747 to 0.809 (*P* value < 0.001). With 100% of labeled data (Supplementary Figure 3), overall Accuracy and F1 score rose further, from 0.844 to 0.873 and from 0.81 to 0.86, respectively, compared with Base BERT (*P* value < 0.001).

Performance was stratified by demographic subgroups (Figs. 2 and Supplementary Figure 3). To evaluate fairness, we applied our PV score, which measures disparities in positive and negative predictive values across groups (see Methods). Lower PV scores reflect more equitable performance.

For race subgroups, with 25% labels (Fig. 2a), Ci-SSGAN achieved the highest Accuracy/F1 across Asian, Black, and White cohorts; its PV was 0.056, lower than Base BERT 0.133 and Bio-Clinical BERT 0.158, but higher than SSGAN 0.027. With 100% labels (Supplementary Figure 3a), Ci-SSGAN again led on Accuracy/F1 and achieved PV = 0.023, matching SSGAN 0.023 and below Base BERT 0.032 and Bio-Clinical BERT 0.077.

For gender subgroups, with 25% labeled data (Fig. 2b), Ci-SSGAN achieved the highest predictive performance (Accuracy/F1) for females (0.830/0.802) and males (0.857/0.809), exceeding SSGAN, Base BERT, and Bio-Clinical BERT. For fairness, Ci-SSGAN was second-best (PV = 0.007), below Base BERT (0.037) and Bio-Clinical BERT (0.066) and slightly above SSGAN (0.003). With 100% labeled data (Supplementary Figure 3b), Ci-SSGAN again led on Accuracy/F1 (female 0.895/0.897; male 0.850/0.840; overall 0.873/0.860) and achieved the lowest PV (0.003 vs SSGAN 0.014, Base BERT 0.020, Bio-Clinical BERT 0.028).

Across age strata (years), Ci-SSGAN consistently achieved the strongest predictive performance. With 25% labeled data (Fig. 2c), accuracy/F1 were 0.836/0.707 for 30–55 y, 0.844/0.817 for 55–70 y, and 0.843/0.807 for ≥70 y; corresponding values were 0.784/0.700, 0.794/0.786, and 0.792/0.748 for SSGAN; 0.789/0.674, 0.749/0.656, and 0.790/0.765 for Base BERT; and 0.787/0.649, 0.730/0.689, and 0.749/0.720 for Bio-Clinical BERT. Fairness, assessed by the PV score, also favored Ci-SSGAN at 25% (PV = 0.025 vs SSGAN 0.063, Bio-Clinical BERT 0.088, Base BERT 0.211). With 100% labels (Supplementary Figure 3c), Ci-SSGAN remained superior on accuracy and F1 across all age groups; its PV was 0.041, lower than Bio-Clinical BERT (0.095) and SSGAN (0.075) and slightly higher than Base BERT (0.035).

Class-wise performance evaluation of the model using PR and ROC analyses demonstrated that Ci-SSGAN achieved the highest overall AUC-PR (0.893), despite an AUC-ROC of only 0.001 below Base BERT (Fig. 3). Ci-SSGAN demonstrated superior precision and recall across most classes, with notably high AUC-PR scores for OAG/S (0.932), ACG/S (0.961), and XFG/S (0.919). For SGL, the rarest class, Ci-SSGAN achieved an AUC-PR of 0.714, substantially outperforming other models. These gains were consistent at 100% labeled data (Supplementary Figure 4).

**Fig. 3.**
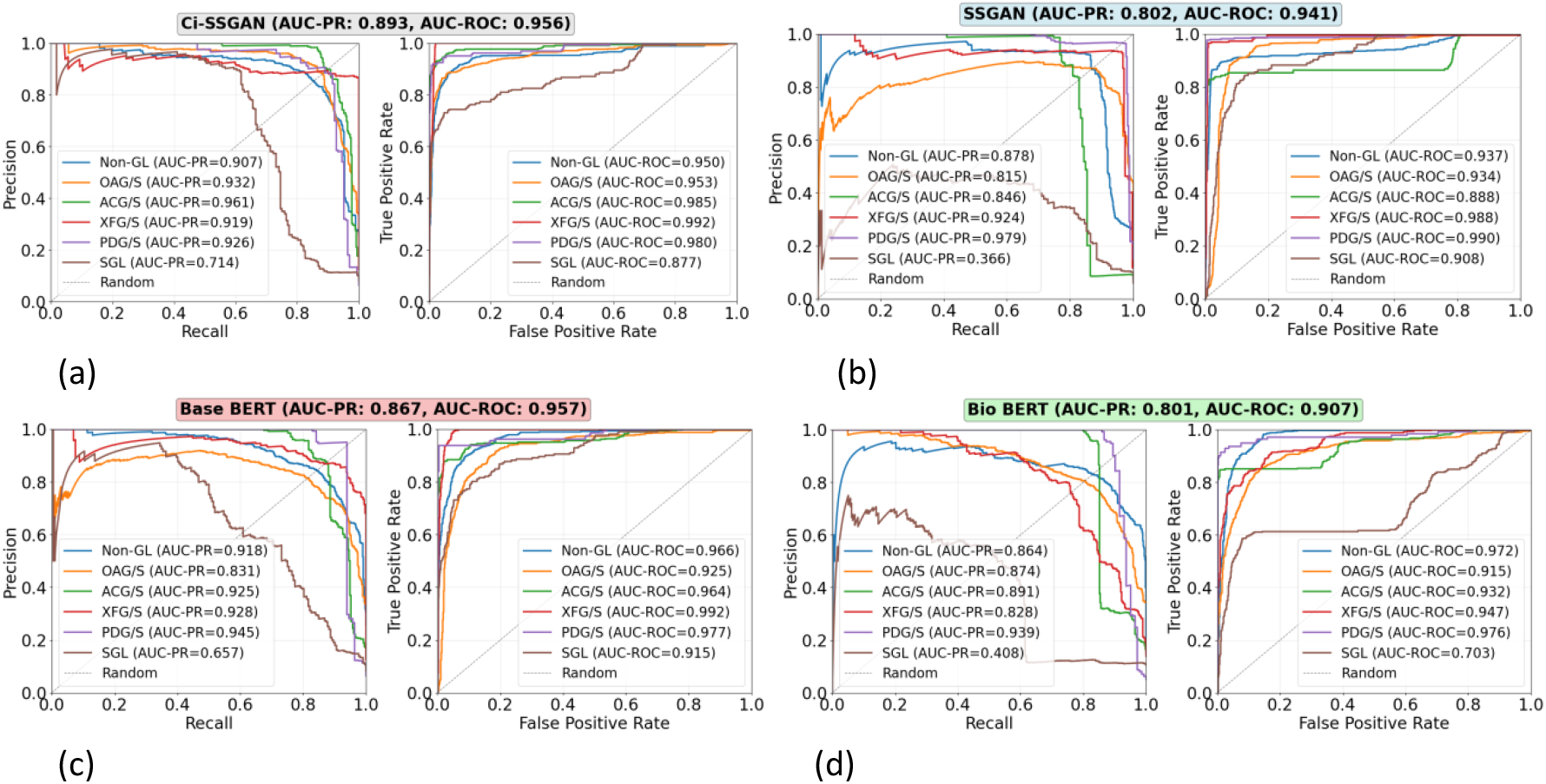
Precision–Recall and ROC curves comparing: **a** Ci-SSGAN, **b** SSGAN, **c** Base BERT, and **d** Bio BERT for multi-class glaucoma subtype and non-glaucoma classification. Each curve shows performance for a specific class, including five glaucoma subtypes and non-glaucoma, with a “Random” baseline for reference showing as dashed line. Ci-SSGAN is designed to condition the generator based on clinical context, while SSGAN is designed without clinical context. Base BERT and Bio BERT are fully supervised baselines. The results are presented on five CV folds. All models were trained using 25% of the labeled data.

### Model interpretability and reliability analysis

To fully characterize models’ behavior, we analyzed feature representation patterns, predictive uncertainty, and token-level interpretability.

The UMAP visualizations in Fig. 4 show that Ci-SSGAN learns more distinct and well-separated patient-level clusters across demographic groups compared to regular SSGAN. Each point represents a patient embedding, derived by averaging note-level feature representations from the discriminator’s final shared layer. Clinically, these clusters correspond to patients with similar disease characteristics, demographic profiles, and glaucoma subtypes, indicating that Ci-SSGAN organizes patients into clinically meaningful groups rather than overlapping latent spaces. Quantitatively, Ci-SSGAN achieved higher silhouette scores for race (0.68), gender (0.72), and age (0.57) compared to SSGAN (0.52, 0.26, and 0.17), reflecting clearer group separation and reduced feature overlap. This improvement is especially pronounced for gender, where Ci-SSGAN’s score (0.72) was significantly higher than SSGAN’s (0.26; *P* value < 0.05), demonstrating that demographic-specific information is better preserved in the learned feature space when clinical context is incorporated.

**Fig. 4.**
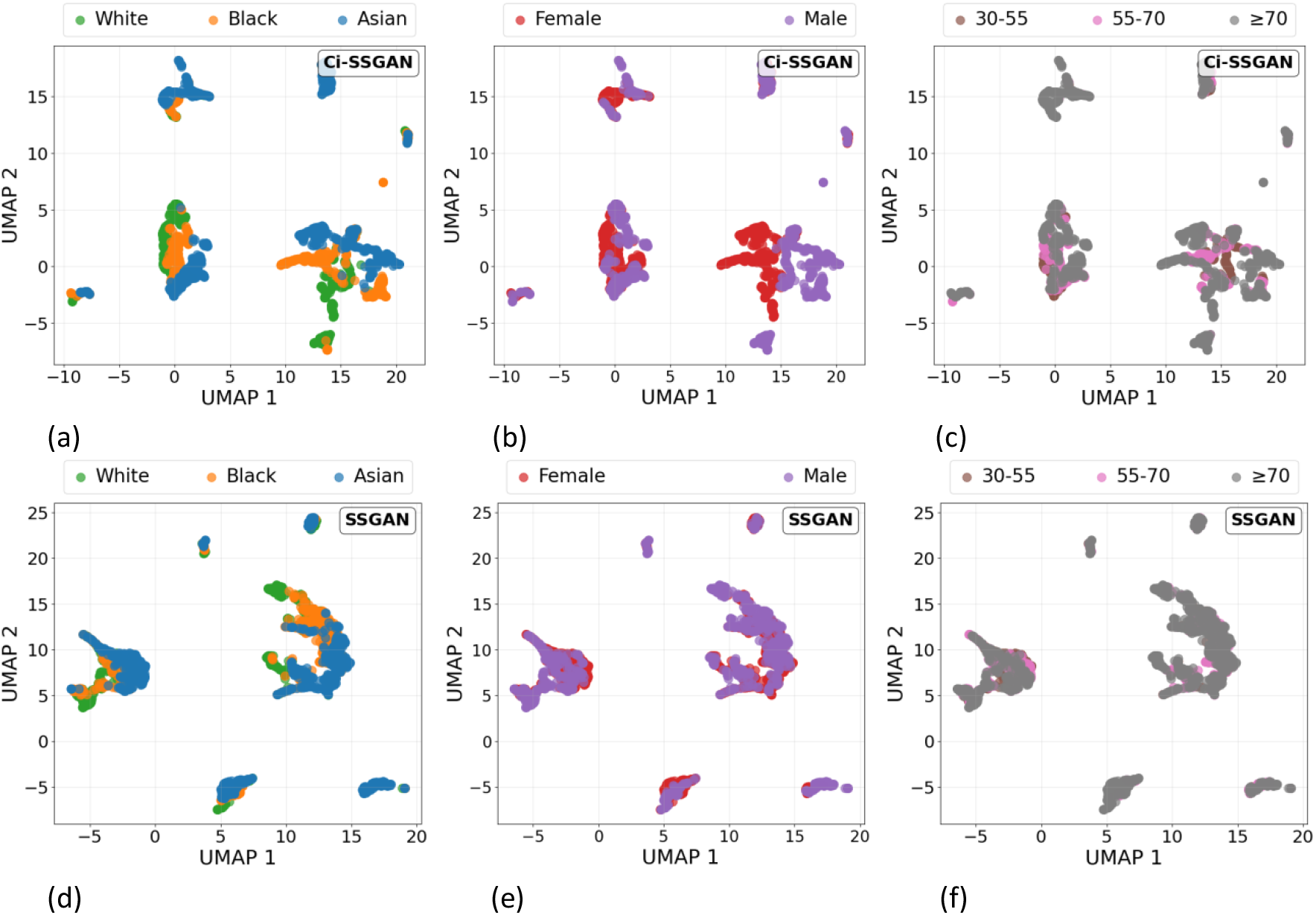
UMAP projections of learned feature embeddings from Ci-SSGAN (top) and SSGAN (bottom), **a, d** colored by race, **b, e** gender, and **c, f** age groups. Each point is a sample, with proximity reflecting similarity in the 256-dimensional discriminator feature space. Ci-SSGAN produces more compact, well-separated clusters across demographics, indicating improved feature organization when incorporating clinical context. Features are from the best fold trained on 25% labeled data.

Across race, gender, and age subgroups, Ci-SSGAN consistently exhibited the lowest predictive uncertainty (Fig. 5), with median entropy values of 0.243 at 25% labeled data and 0.263 at 100% labeled data, compared with SSGAN (0.5 and 0.6), Base BERT (0.727 and 0.458), and Bio BERT (0.687 and 0.447). When increasing the labeled fraction from 25% to 100%, the change in median entropy was minimal for Ci-SSGAN (−5.3%), indicating stable confidence even in low-label regimes, whereas SSGAN showed a 16.9% reduction, Base BERT 34.1%, and Bio BERT 32.0%, reflecting greater dependency on labeled data for uncertainty reduction. This trend was consistent across racial groups (Asian, Black, White), gender groups (male, female), and age ranges (30-55, 55-70, ≥ 70), with the performance gap most pronounced under the 25% labeled data condition. Fig. 5 illustrates these results.

**Fig. 5.**
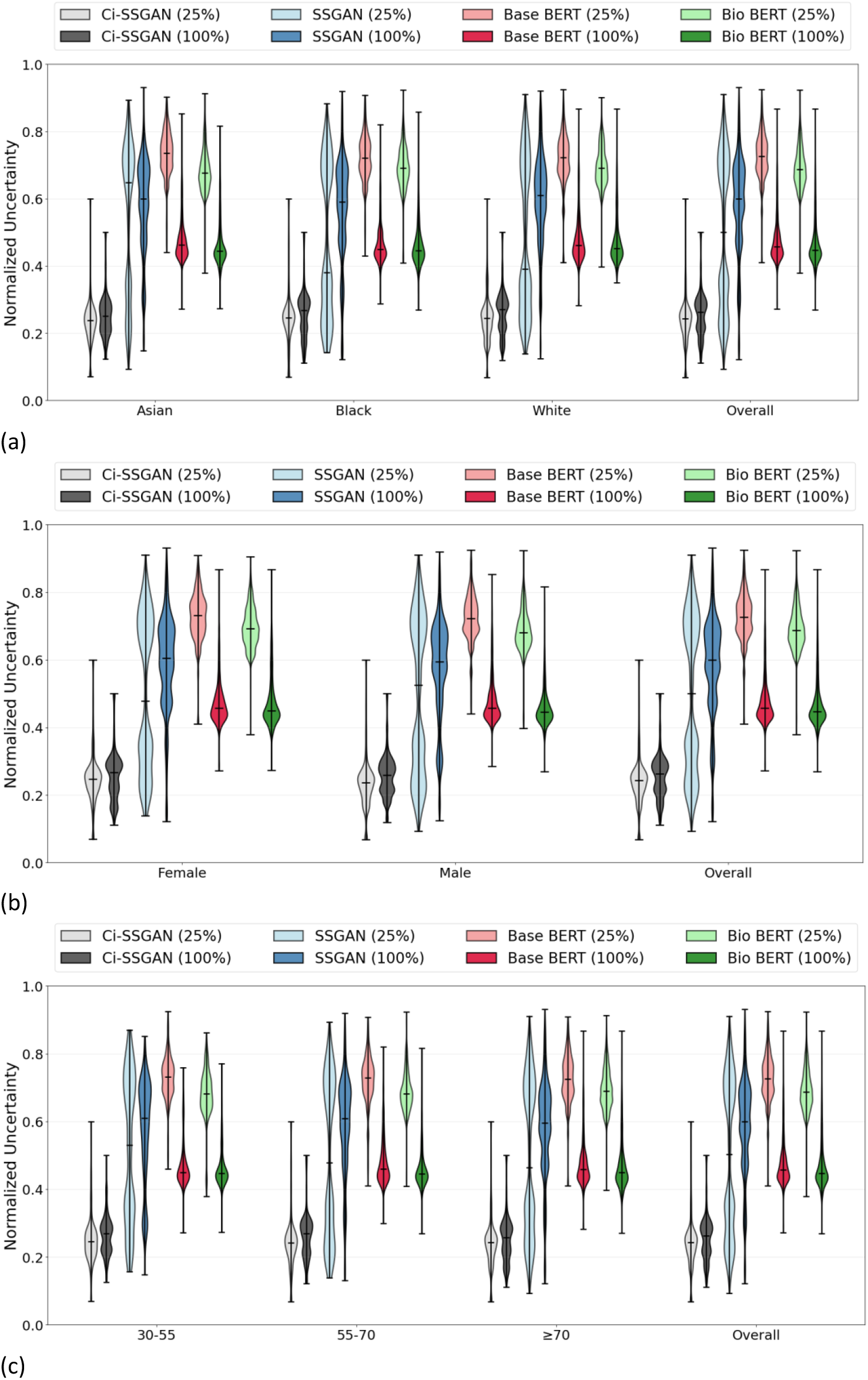
Uncertainty analysis across demographic groups for models trained with 25% and 100% of labeled data. Violin plots show entropy distributions for four models across: **a** race, **b** gender, and **c** age groups. Light colors indicate 25% training data; dark colors indicate 100% data. Ci-SSGAN (gray) maintains lowest uncertainty (< 0.2 entropy) regardless of data size, while BERT models (red/green) show high uncertainty with 25% data (0.6-1.4 entropy) that reduces substantially with 100% data (0.2-0.4 entropy). SSGAN (blue) exhibits intermediate performance. The horizontal line indicates the median and the error bars represent the minimum and maximum values observed for each group.

Clinically, these results indicate that Ci-SSGAN not only improves classification performance but also delivers markedly more reliable predictions. Supplementary Figure 5 supports this by showing class-wise probability radar plots where Ci-SSGAN predictions form tightly clustered, high-confidence outputs (mean confidence = 0.990, ACC = 0.890) compared with the more dispersed, less certain outputs of standard SSGAN (mean confidence = 0.975, ACC = 0.829).

Gradient-weighted token attribution, averaged across five folds, shows model-specific patterns of feature use (Fig. 6). For display, we first select the top 20 tokens per model after HIPAA filtering and then renormalize their scores to sum to 1.0 within each model. Under this convention, Ci-SSGAN exhibits a concentrated profile, with the wordpiece “igmentary” (from pigmentary glaucoma) accounting for 0.478 of the displayed attribution, followed by “glaucoma” (0.102) and “pseudoexfoliation” (0.019), consistent with clinically focused conditioning. SSGAN is more diffuse (maximum 0.195 for “fovea”), Base BERT the most dispersed (maximum 0.103 for “cyclophotocoagulation”), and Bio-Clinical BERT similarly diffuse (maximum 0.073). The maximum displayed attribution share is 6.5-fold higher for Ci-SSGAN than for Bio-Clinical BERT (0.478 vs 0.073), suggesting more concentrated, disease-oriented token selection with architectural clinical conditioning. Because the plot reflects relative attribution among the top tokens aggregated across notes, it should be interpreted as a comparative profile rather than evidence of token frequency; per-class token profiles are provided in Supplementary Figure 6.

**Fig. 6.**
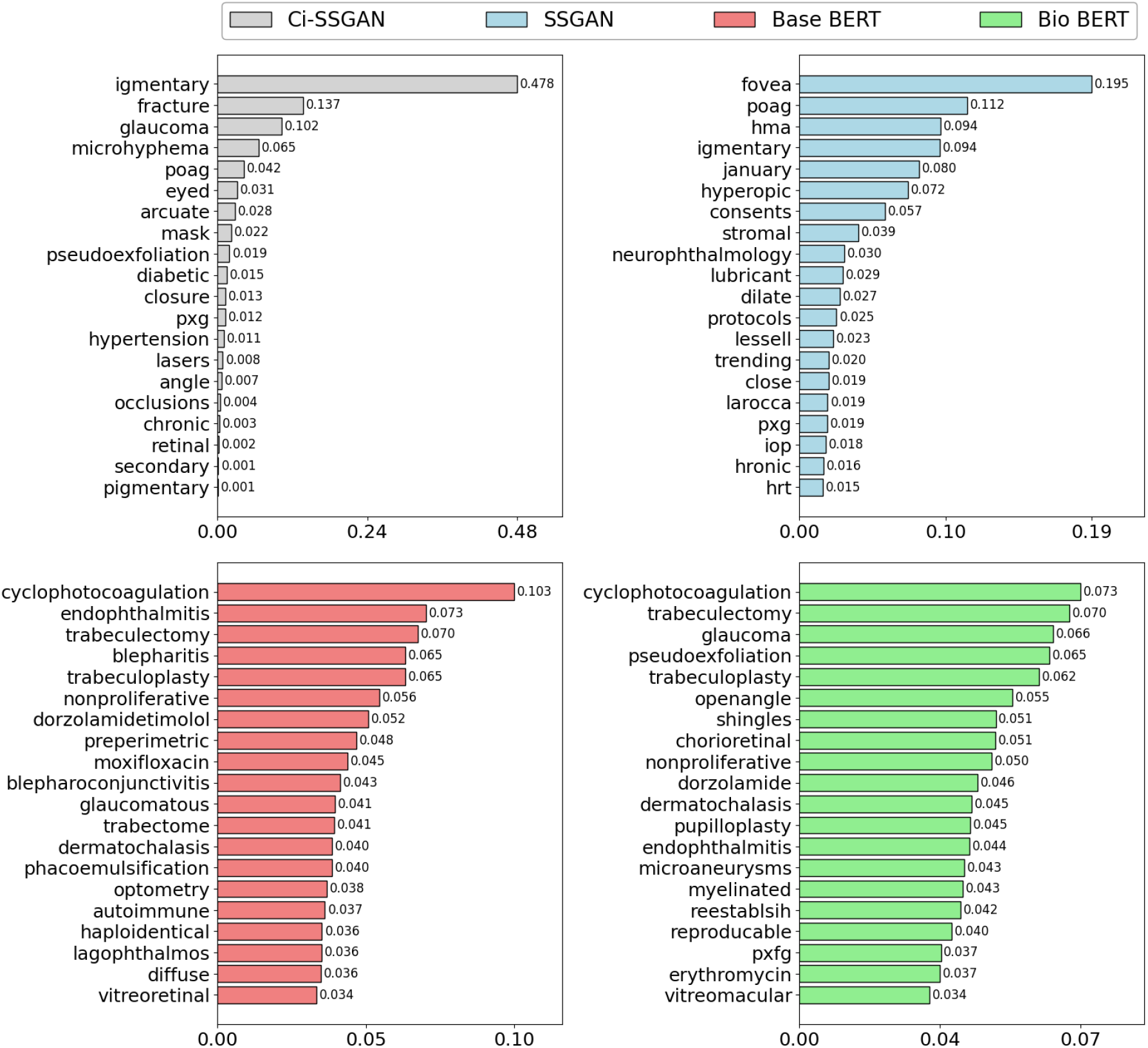
Gradient-weighted token attribution by model. Wordpiece-level attributions are normalized per note to sum to 1 and then averaged across notes (5-fold CV). To avoid rare-token artefacts, only tokens that appear in ≥ 50 notes (≥100 total occurrences) are considered. Bars show the mean normalized attribution share for the top 20 tokens per model. Values printed on the bars represent the distribution of attribution mass among the displayed top tokens, not corpus-level frequency. Models trained by 25% of labeled data were used to extract these tokens.

### Prediction Performance of Ci-SSGAN subtype detection Versus ICD Code labels on the Test Dataset

We further demonstrate that our model can outperform ICD code-based labels, using glaucoma subtype detection as a test case which suffer from coarse and noisy ICD codes, frequently requiring re-annotation. Ci-SSGAN yielded confusion matrices with reduced cross-subtype misclassifications and closer alignment with expert clinical grading compared with ICD-based labels (Fig. 7). Of the 105 test patients, 94 had ≥1 ICD code that mapped unambiguously to a glaucoma subtype; the remaining 11 lacked a mapped code or had only non-specific/suspect codes and were excluded from the ICD confusion matrix.

**Fig. 7.**
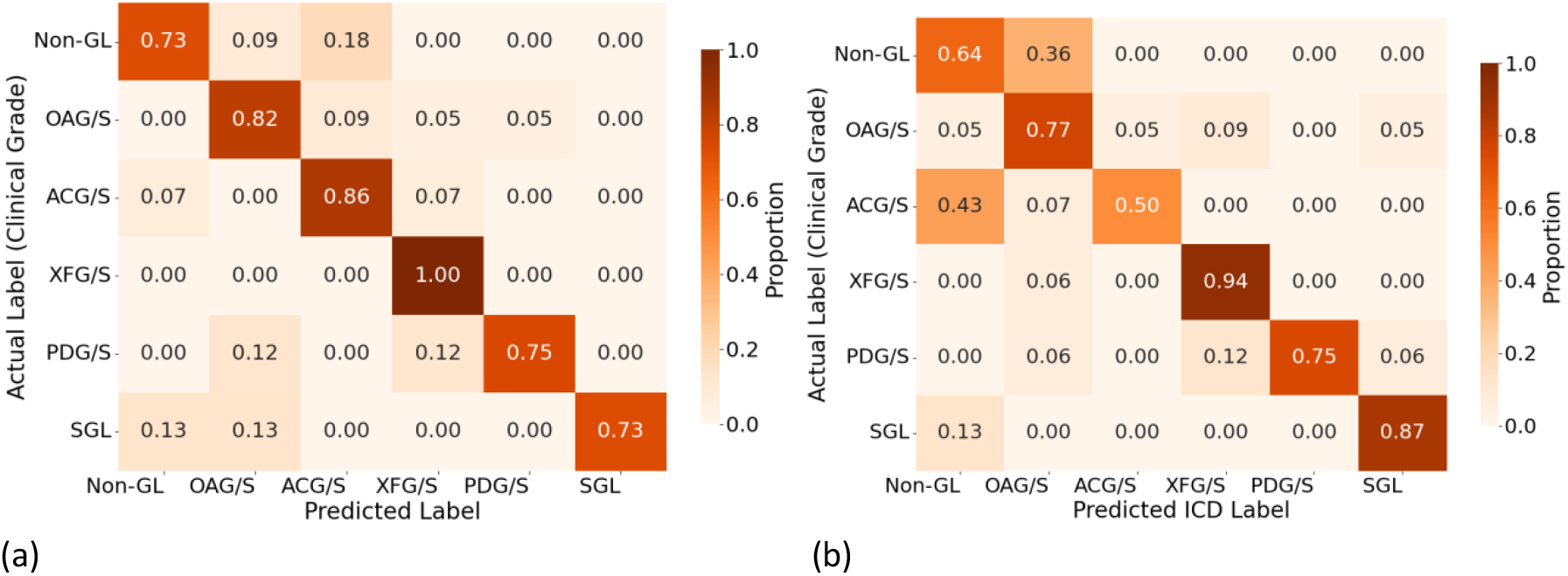
Confusion matrices comparing Ci-SSGAN and ICD-based labels against expert clinical grading for glaucoma subtypes. **a** Ci-SSGAN predictions derived from free-text clinical notes. **b** ICD-based subtype assignments derived from structured billing codes. Ci-SSGAN demonstrates fewer cross-subtype misclassifications and stronger concordance with expert labels across all six subtypes: Non-GL, OAG/S, ACG/S, XFG/S, PDG/S, and SGL. ICD-based labels were assigned using predefined diagnostic code mappings, with a prioritization hierarchy applied when multiple codes were present. Of 105 patients in the test cohort, 94 had at least one valid ICD code. The remaining 11 had no linked billing record or only non-specific/suspect codes (e.g., H40.9/365.9), which we do not use for subtype assignment.

Ci-SSGAN demonstrated superior performance over ICD-based labels across all evaluation metrics (Fig. 8 and Supplementary Figure 7). Overall accuracy was significantly higher for Ci-SSGAN (0.853) compared with ICD-based labels (0.744, *P* value < 0.05), with substantial per-class improvements in Non-GL (0.818 vs. 0.636, *P* value < 0.05), OAG/S (0.955 vs. 0.773, *P* value < 0.05), and ACG/S (0.929 vs. 0.500, *P* value < 0.05). Similarly, the overall F1 score was greater for Ci-SSGAN (0.855 vs. 0.744, *P* value < 0.05), with the largest margins in Non-GL (0.750 vs. 0.519, *P* value < 0.05), OAG/S (0.857 vs. 0.739, *P* value < 0.05), and ACG/S (0.929 vs. 0.636, *P* value < 0.05).

**Fig. 8.**
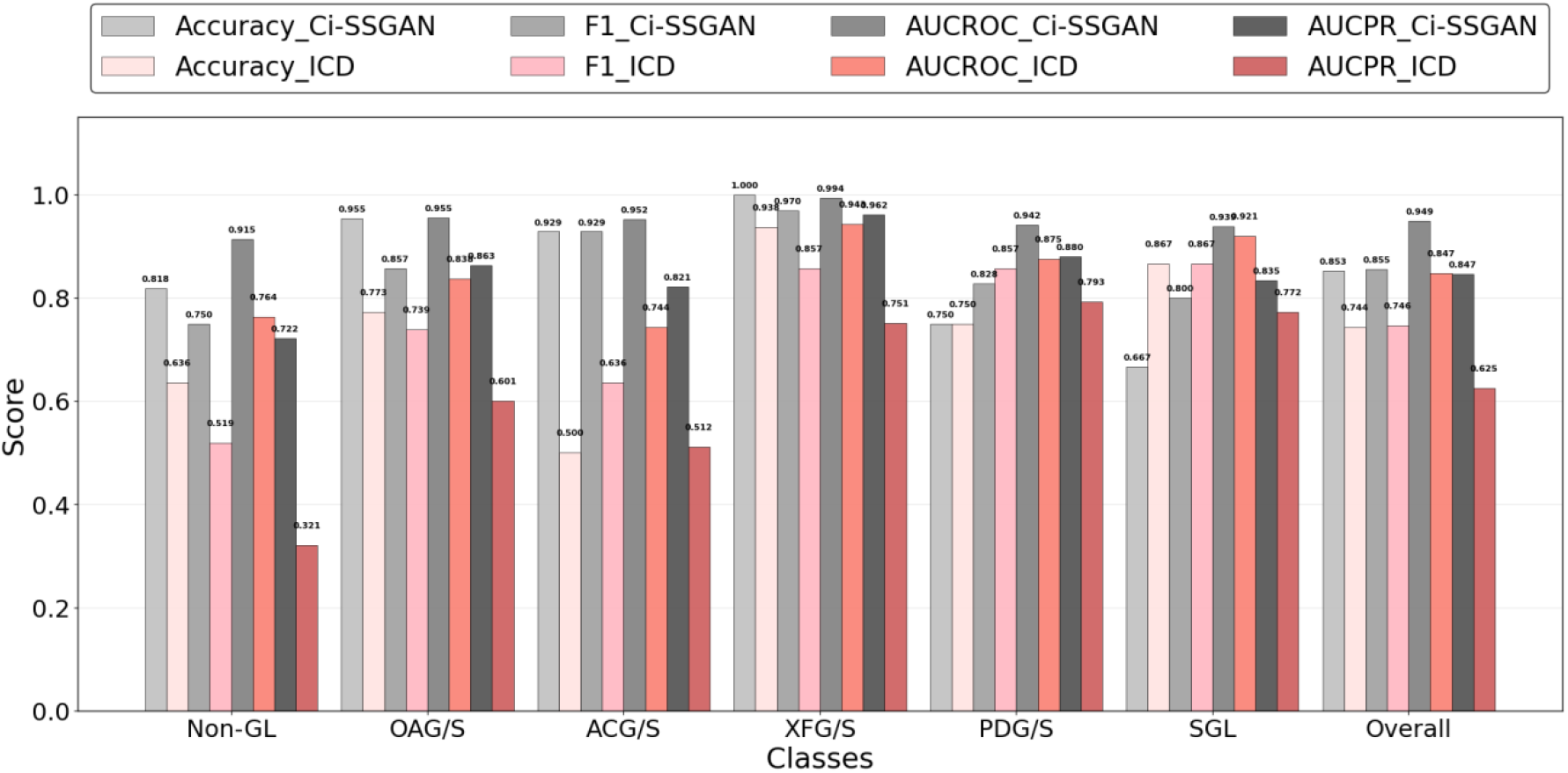
Comparison of Ci-SSGAN and ICD code–based labeling across glaucoma subtypes and non-glaucoma cases in terms of Accuracy, F1 Score, AUC-ROC, and AUC-PR. Each bar represents performance for a specific class, with the “Overall” category summarizing all classes. Ci-SSGAN uses both labeled and unlabeled data with clinical context, whereas ICD code labels rely solely on diagnosis codes from medical records. Improvements were most pronounced in challenging subtypes such as primary angle-closure glaucoma (ACG/S), open-angle glaucoma (OAG/S), and Non-GL cases.

From a clinical standpoint, the improvements in AUROC and AUPRC are substantial. Relative to ICD, Ci-SSGAN achieved a higher AUROC (0.949 vs 0.847, *P* value < 0.01), with per-class gains in Non-GL (+0.151), OAG/S (+0.117), and ACG/S (+0.208) (*P* value < 0.05 each). AUPRC (which is sensitive to class prevalence and emphasizes precision at high recall) also improved overall (0.847 vs 0.619, *P* value < 0.01). The largest per-class AUPRC gains were observed for Non-GL (+0.401, indicating fewer false positives across glaucoma subtypes), OAG/S (+0.262), and ACG/S (+0.309) (*P* value < 0.05 each). Although AUPRC is particularly informative for rarer subtypes, we report it for all classes, including the majority Non-GL class, to provide a complete per-class view. Overall performance metrics for all models are summarized in Table 3.

**Table 3.** Overall performance summary of Ci-SSGAN vs. ICD code. All values are calculated on the test dataset. Bold values show the best scores.

| Metric | ICD Code | Ci-SSGAN (25%) | Ci-SSGAN (100%) | Gain vs. ICD (25%) | Gain vs. ICD (100%) |
| --- | --- | --- | --- | --- | --- |
| Accuracy | 0.743 | **0.773** | **0.853** | +2.96 | +10.91 |
| F1 score | 0.746 | **0.765** | **0.856** | +1.93 | +10.97 |
| AUROC | 0.847 | **0.942** | **0.949** | +9.46 | +10.19 |
| AUCPR | 0.625 | **0.821** | **0.847** | +19.62 | +22.22 |

## Discussion

This study demonstrates that clinically informed semi-supervised generative modeling can address two long-standing challenges in healthcare AI: the scarcity of reliable labels and the inequities in subgroup performance, by harnessing abundant unlabeled clinical notes. Glaucoma exemplifies the urgency of these challenges and the potential of Ci-SSGAN to mitigate them, particularly for under-represented groups such as Black patients, females, and younger individuals who experience worse disease outcomes ^22,23,28,29^. Biases in structured EHR labels, especially when combined with limited annotations, have been shown to undermine model reliability ^11,14,15^, and prior work highlights how feature-dependent noise, annotator disagreement, and limited diversity in training corpora degrade performance in ways that fine-tuning alone cannot resolve ^16,17^. While semi-supervised GANs have attempted to address data scarcity, they typically depend on noise or broad demographic inputs ^18,19,30,31^, reducing their ability to capture clinically meaningful signals. Unlike standard SSGAN models used in image classification ^18,19,31^, where the generator is conditioned only on noise or limited demographics, Ci-SSGAN introduces three key innovations: integration of note embeddings to capture clinical context, multi-conditional learning to counter subgroup imbalance, and an explicit fairness objective using the PV metric.

Across varying amounts of labeled data, Ci-SSGAN consistently outperformed both supervised and semi-supervised baseline models (Table 2 and Figs. 2 and 3). Its conditional generation on clinical embeddings enabled robust improvements even with limited annotations, and these benefits extended to high-label settings where other models typically plateau. For example, with only 25% labeled data, Ci-SSGAN achieved an accuracy of 0.871 ± 0.03, surpassing SSGAN (0.859 ± 0.04) and Base BERT (0.824 ± 0.05), alongside a substantial AUC-PR improvement (+0.091 over BioBERT). These advantages persisted as labeled data increased with the framework achieving over 9% higher precision–recall performance compared to standard supervised approaches, illustrating its ability to extract value from unlabeled notes while maintaining equitable subgroup performance.

Importantly, Ci-SSGAN semi-supervised design enables remarkable label efficiency. Comparing 25% to 100% labeled data (Supplementary Table 1), performance gains were modest (accuracy +0.036, F1 +0.039, AUROC +0.007, AUCPR +0.026), indicating that the model extracts near-optimal information from unlabeled notes. This efficiency lowers the barrier to AI development, enabling strong results without exhaustive annotations, which is particularly valuable for rare diseases, underrepresented groups or when resources are limited. Notably, improvements remained substantial even with 100% labeled data (Table 3), not contradicting the label-efficiency advantage but highlighting complementary effects: in low-label regimes, Ci-SSGAN leverages unlabeled notes to compensate for limited supervision, while in high-label regimes it combines full supervised signal with adversarially generated diversity, pushing beyond the ceiling of purely supervised models. While BERT variants plateau once additional labels no longer reduce uncertainty, Ci-SSGAN continues to improve by preserving rare subtypes and demographic variability— explaining why its margin of improvement can appear larger at 100% labeled data, even though relative efficiency is most impactful with limited data.

Ci-SSGAN also produced more discriminative feature representations and reduced predictive uncertainty (see Supplementary Figure 5), supporting reliable outputs across demographic subgroups. Attribution analysis further showed that the model concentrated attention on disease-specific terms (e.g., “pigmentary,” “pseudoexfoliation”), with over six-fold greater attribution focus compared to Bio BERT. This sharper reliance on clinically meaningful features provides mechanistic insight into why the framework achieves both superior performance and interpretability. Importantly, while BERT models became increasingly confident only as more labels were added, Ci-SSGAN maintained the lowest overall uncertainty and preserved sensitivity to rare subtypes. This balance between certainty and diversity prevents overconfident errors and underscores the framework’s design to promote equitable performance.

When benchmarked against ICD-based glaucoma coding, Ci-SSGAN showed markedly higher concordance with expert grading (Table 3 and Fig. 8), particularly for open-angle and angle-closure subtypes and in distinguishing non-glaucoma cases. These results highlight how leveraging unlabeled notes can overcome the coarse granularity and systematic misclassification inherent to structured coding. From an equity perspective, PV scores were substantially reduced across race, gender, and age, with notable gains for Black patients (+6.7% F1 vs. Bio BERT), illustrating that conditioning generative models on unlabeled clinical text offers a viable path to mitigating potential systemic biases.

Although our case study focused on glaucoma, the architecture is conceptually applicable to other medical conditions with abundant clinical documentation but limited labels, such as diabetic retinopathy staging ^32-34^, cardiac disease risk stratification ^35-37^, or psychiatric disorder classification ^38-40^. The main limitation preventing validation in these areas is the lack of large-scale, expert-annotated note datasets, which constrains systematic evaluation beyond glaucoma. While further validation will be needed once such resources become available, our results suggest that multi-conditional semi-supervised learning on unstructured text has the potential to improve accuracy, reliability, and fairness in diverse healthcare contexts.

We explicitly acknowledge several limitations we faced in this study. Data were drawn from a single academic health system; external validation is required to ensure generalizability across institutions and note types. Smaller demographic groups (Hispanic/Latinx, Indigenous, multi-racial) were underpowered and should be included in future equity analyses. Expert annotations, while highly consistent (kappa ≈ 0.90), may reflect prevailing clinical biases. From a translational perspective, future deployment of Ci-SSGAN would likely require integration into clinical decision-support systems (CDS) with careful evaluation of human–AI collaboration strategies (e.g., clinician override, uncertainty visualization, and workflow co-design), to ensure safe and equitable use in practice.

Building on these considerations, future work should include multi-center validation to ensure robustness across diverse populations, and prospective clinical evaluation to measure outcomes such as time-to-diagnosis and vision preservation. In addition to methodological safeguards such as differential-privacy training and threat modeling, improved note-derived annotations could directly enable construction of more accurate cohorts for large-scale epidemiologic and genetic studies, as well as better-powered clinical trials. Integration into CDS systems could facilitate more accurate patient labeling, risk stratification, and referral triaging, thereby enhancing early detection and timely care. Comprehensive fairness audits, including intersectional subgroup analyses and debiasing strategies, remain essential to address residual disparities. Finally, co-designed workflows with clinicians and patients, particularly in under-resourced communities, will be key to translating technical advances into equitable care delivery.

In conclusion, this study demonstrates that semi-supervised generative model, when grounded in clinical context and demographic information, can simultaneously improve accuracy and equity in healthcare AI. By leveraging abundant unlabeled notes, Ci-SSGAN reduces dependence on large, annotated datasets and maintains subgroup performance that typically degrades in low-data settings. These findings highlight the broader potential of clinically informed semi-supervised learning to address both data scarcity and systemic bias in real-world applications.

## Methods

The Institutional Review Board at Mass General Brigham (MGB) approved this study, which adhered to the ethical guidelines outlined in the Declaration of Helsinki for research involving human participants. Given the retrospective design, the requirement for informed consent was waived.

### Inclusion and exclusion criteria

In our MEE dataset, which encompasses over 250 million notes, we included notes from the ophthalmology departments comprising 3.2% of total notes. We filtered the notes with dates after May 2015 and up to December 2024. The majority of physician-generated notes are concentrated in two primary categories: “Progress Note” and “Assessment and Plan Note,” collectively totaling over 90 million notes. The notes vary in length with min and max lengths of 1 and 386561 characters, respectively (Interquartile range = 455-1672 characters. For the analysis and to account for system limitations, we limited the note length between 200 to 5000 characters, approximately 40 to 1100 tokens. A sliding window approach was used for longer notes exceeding the tokenizer’s maximum length of 512 tokens to split them into overlapping chunks of 512 tokens (overlap=64) to ensure context continuity. We specifically included only White or Caucasian, Black or African American, and Asian patients due to the abundance of notes in our dataset. The final dataset comprises 2129171 notes associated with 327814 patients, consisting of 88.2% White, 6.8% Black African Americans, and 5% Asians. Supplementary Figure 8 shows the department specialties, note type, and note length in our dataset.

### Manual review and labeling

Notes were initially filtered by glaucoma-related keywords extracted from the literature and online ophthalmology resources ^41^. For each condition, at least 50 patients with a minimum of two notes were randomly selected. Six independent clinicians reviewed the filtered notes for five glaucomatous conditions—open-angle glaucoma/suspect (OAG/S; including normal-tension glaucoma, high-tension glaucoma, and ocular hypertension), angle-closure glaucoma/suspect (ACG/S), exfoliation glaucoma (XFG), pigmentary dispersion glaucoma (PDG), and secondary glaucoma (SGL)—as well as related suspect/syndrome categories (open-angle suspect [OAS], angle-closure suspect [ACS], exfoliation syndrome [XFS], pigment dispersion syndrome [PDS]), ensuring that each patient had notes on at least two dates. Inter-rater agreement on a 10% subset yielded Cohen’s kappa (*κ*)=0.90, indicating excellent concordance. Notes without glaucomatous signs were labeled Non-GL; uncertain notes were labeled Other and excluded from training. The final curated set comprised 3035 notes from 1282 patients and served as the ground truth for the supervised portion of model training.

Structured labels were also derived from ICD-10/ICD-9 billing codes (pre- and post-October 2015) to construct an ICD-based baseline. Mapping was: XFG/S: H40.14, 365.52; PDG/S: H40.13, 365.13; OAG/S (including suspects and ocular hypertension): H40.1, H40.01, H40.02, H40.05, 365.1, 365.01, 365.04, 365.05; ACG/S: H40.2, H40.03, H40.06, 365.2, 365.02, 365.06; SGL: H40.3, H40.4, H40.5, H50.6, 365.3, 365.4, 365.5, 365.6. General or nonspecific glaucoma codes were mapped to Non-GL. For patients with multiple subtype codes, a predefined priority (XFG/S > PDG/S > SGL > ACG/S > OAG/S > Non-GL) was applied to assign a single label, following our prior mapping strategy ^42^. These ICD-derived labels were used only for the ICD-based baselines and for comparison with Ci-SSGAN. The workflow for dataset generation, including data collection, preprocessing, and labeling, is shown in Supplementary Figure 9.

### Data preprocessing

All notes were de-identified using the Philter package to ensure HIPAA compliance ^43^. Text was tokenized with BioClinical BERT ^2,3^, and processed using a sliding window strategy to handle the tokenizer’s 512-token limit. Notes longer than 512 tokens were split into overlapping segments of 512 tokens (with 64-token overlap) to preserve context continuity. Each segment was padded if shorter than 512 tokens and wrapped with [CLS] and [SEP] tokens. Demographics (age, gender, race) were encoded numerically, with age normalized to 0-1 and categorical variables converted to integer representations.

For model training, the labeled dataset was split into 90% for training and validation (1000 patients, 2660 notes) and 10% for a held-out test set (105 patients, 294 notes). Patient-level splits ensured that notes from the same individual did not appear in both training and evaluation sets. Five-fold cross-validation was used within the training partition, with a further 10% of patients in each fold set aside for early stopping.

To increase the diversity of training samples, we implemented a custom text augmentation pipeline for both labeled (validation and test sets were not augmented) and unlabeled notes. Augmentation (shuffling, synonym substitution, abbreviation expansion, and full spelling) operated at the note level, generating multiple synthetic variants per original note. Specifically, our pipeline used randomized substitutions and structural edits to mimic clinical note variability while preserving semantic meaning. The augmentation rates adjusted to balance race–class distributions. After augmentation, the training corpus increased from 2660 to 12825 notes. For the unlabeled partition, the corpus increased from 349587 to 1399568 notes. The data preprocessing, and augmentation steps are illustrated in Fig 9.

**Fig. 9.**
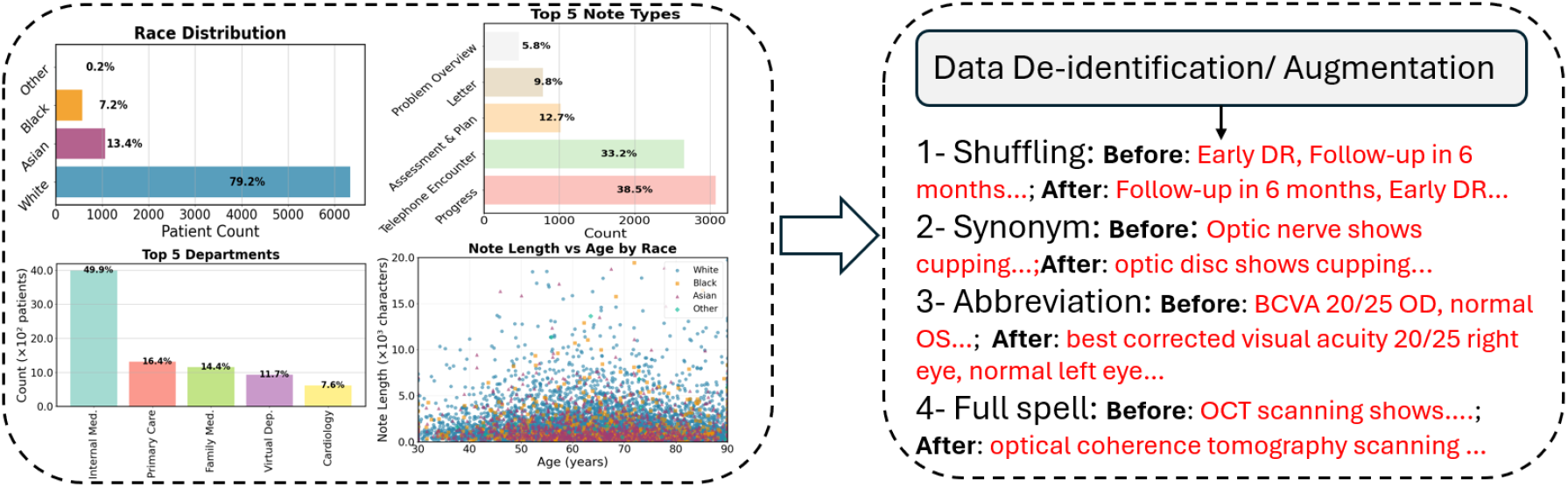
Overview of dataset characteristics and preprocessing pipeline. Patient demographics, note types, and clinical note distributions are shown on the left. Processed notes are then de-identified and augmented (token shuffling, synonym substitution, abbreviation expansion, and full spelling), as illustrated on the right.

### The proposed Ci-SSGAN architecture

Unlike regular SSGANs which only use random noise as input to the generator^18,30,31,44,45^, Ci-SSGAN introduces a novel architecture where the generator directly learns from unlabeled clinical text. The model comprises three components: (1) a Bio-ClinicalBERT text encoder producing 768-dimensional embeddings from clinical notes; (2) a clinically-informed generator accepting 871-dimensional input, combining 100-dimensional Gaussian noise, 3-dimensional demographics (age, race, gender), and crucially, 768-dimensional unlabeled text embeddings; and (3) a dual-head discriminator with shared feature extraction for both 6-class glaucoma classification and real/fake discrimination. This architecture enables the model to learn medical language patterns from large unlabeled clinical notes to improve classification accuracy when labeled data are scarce. All BioClinicalBERT parameters were fine-tuned (no freezing was applied), so both the transformer weights and the fully connected layers were updated during training. The generator, discriminator, and text encoder were trained jointly in an adversarial framework using separate optimizers and hyperparameters with a multi-component loss combining supervised focal loss, adversarial loss, feature matching, clinical consistency, and diversity terms to address class imbalance, preserve clinical semantics, and prevent mode collapse. A detailed description of the model learning curves, model architecture, and full list of loss functions are provided in the Figures 10 and 11 and Tables 2 and 3 in the Supplementary material. An overview of the proposed Ci-SSGAN architecture, including the generator and discriminator components with their inputs, is shown in Fig. 10.

**Fig. 10.**
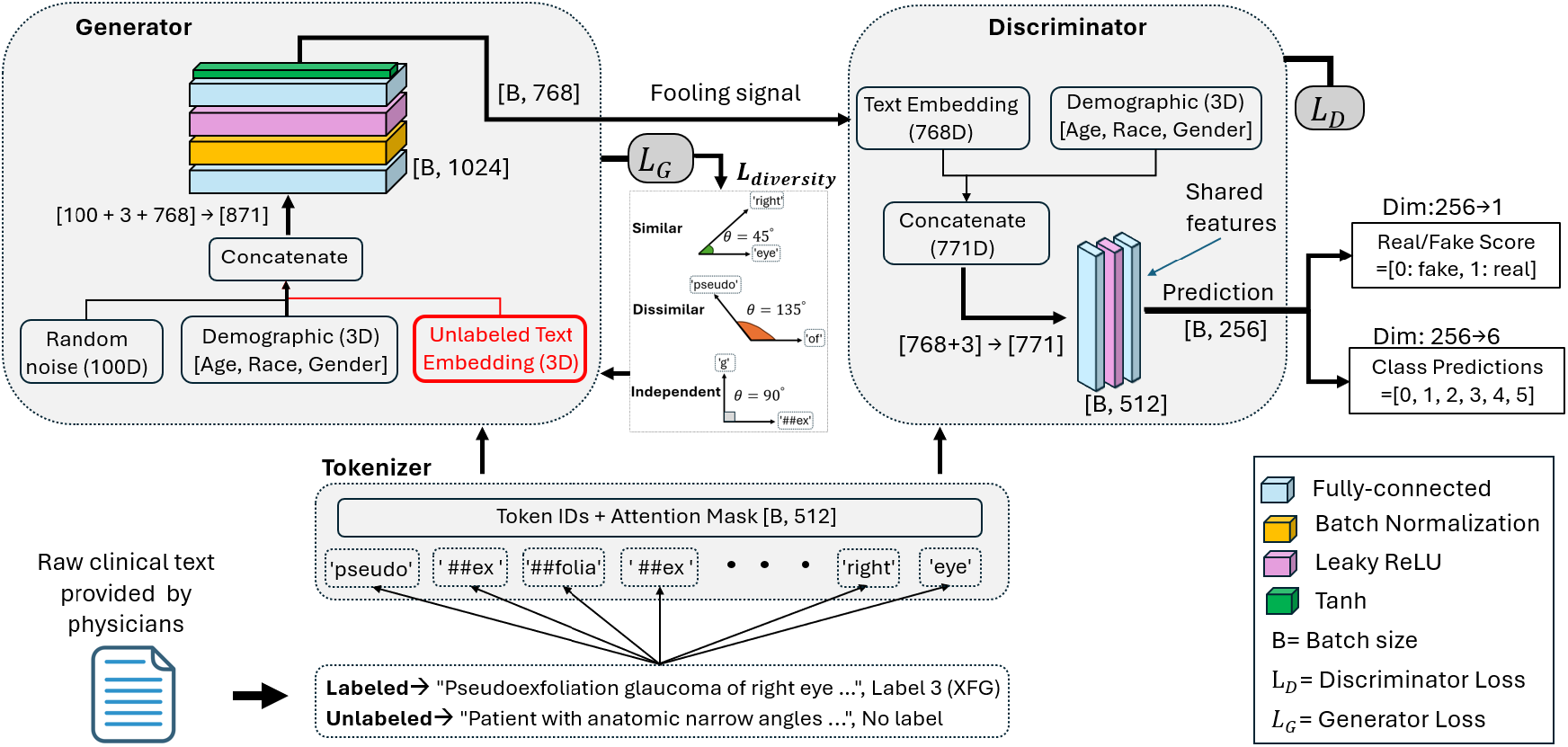
Architecture of the proposed Ci-SSGAN model. The generator takes as input random noise, demographic features (age, race, gender), and embeddings from unlabeled clinical text, producing synthetic text embeddings to fool the discriminator. The discriminator processes both real and generated embeddings along with demographic features, outputting a real/fake score and class predictions. The labels 0-5 refer to: Non-GL, OAG/S, ACG/S, XFG/S, PDG/S, and SGL, respectively. Demographics make the generator demographically conditioned synthesis as it knows how to create appropriate synthetic data for each type of patient. For clarity, only diversity loss is shown. *L*_G_ is also trained with adversarial, feature matching, and clinical consistency losses. All components, including the BioClinicalBERT encoder and fully connected layers, were fine-tuned. Generator and discriminator were updated alternately using separate optimizers and hyperparameters.

### Training protocol

Training was conducted using Python 3.10 and PyTorch 2.3.1 on eight NVIDIA H100 GPUs 80 GB memory each) in a distributed data-parallel setup. Hyperparameters, including learning rates for each model component, were optimized using Optuna ^46^ with 50 trials. The final selected learning rates were 3 × 10^−5^ for the text encoder, 1 × 10^−4^ for the discriminator, and 2 × 10^−4^ for the generator. Models were trained for up to 250 epochs with early stopping (patience = 11), batch size = 16, and gradient clipping at a max norm of 0.5. Underrepresented classes, particularly SGL, were upweighted using dynamically calculated class weights with a 5× boost factor. Focal Loss (*γ* = 2) was used to emphasize hard-to-classify cases. The semi-supervised training framework incorporated 349853 unlabeled notes from 155392 patients, utilized in both standard SSGAN and Ci-SSGAN training. Unlabeled embeddings were provided to the generator alongside noise and demographics, enabling clinically relevant synthetic sample generation and enhancing feature diversity. Model comparisons were performed against baseline models (Base BERT and BioClinical BERT) trained with full parameter fine-tuning. Supplementary Table 4 shows the full list of hyperparameters and model configurations.

### Performance metrics and statistical analysis

Model performance was evaluated using accuracy, F1 score, area under the receiver operating characteristic curve (AUROC), and area under the precision–recall curve (AUCPR). These metrics assessed both overall glaucoma detection accuracy and demographic-specific performance across racial, gender, and age subgroups, with analysis conducted at both note and patient levels. Accuracy provides a global measure of correct classifications, while the F1 score balances precision and recall, offering a clinically meaningful view of how well the model detects glaucoma cases without excessive false positives. AUROC measures the model’s ability to discriminate between disease and non-disease across all decision thresholds. AUCPR emphasizes precision in identifying true positive cases. We evaluated performance stratified by age groups (30-55, 55-70, >=70 years), gender (female, male), and race (Asian, Black, White). To quantify model’s bias across demographics, we introduce a new *PV* score that simultaneously evaluates over-diagnosis and under-diagnosis disparities across demographic Categories (*C*) using *PPV* and *NPV*. This metric bridges model’s fairness with clinical practice by measuring disparities in both false positive and false negative prediction rates and is formulated as:

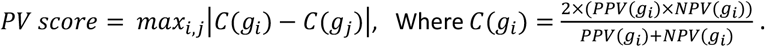

Here, *g*_i_ and *g*_j_ are individual groups within that category. This formulation bridges fairness with clinical practice by penalizing disparities in both false-positive and false-negative rates. Prediction uncertainty for each sample was determined using Tsallis entropy ^47^, computed as

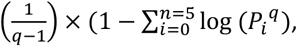

where *P*_i_ is the predicted probability of class *i*. Given the class imbalance in glaucoma subtype classification, we set the entropic parameter to *q* = 0.25, which emphasizes uncertainty in rare classes and provides greater sensitivity to predictions involving underrepresented glaucoma types ^48^.

All tests were two-sided with significance set at *P* < 0.05. Model performance was compared across metrics, data fractions, and demographic subgroups. Pairwise differences (accuracy, F1, AUROC, AUCPR, PV score) were assessed using paired t-tests across five cross-validation folds. Subgroup analyses (race, gender, age) used ANOVA with post-hoc t-tests. Multiple comparisons were adjusted with Benjamini– Hochberg false discovery rate (FDR) correction, and adjusted *P* < 0.05 was considered significant. Analyses were performed in Python 3.10 using SciPy (v1.13) and statsmodels (v0.14).

## Data availability

The data can be obtained from the corresponding author upon request.

## Code availability

The codes for the proposed model are available at https://github.com/Mousamoradi/Ci-SSGAN.

## Acknowledgments

This research was funded by the National Institutes of Health (NIH: R01 EY036222 and R21 EY035298), and MIT-MGB AI Cures Grant.

