## Supplementary File for "Clinically Informed Semi-Supervised Learning Improves Disease Annotation and Equity from Electronic Health Records: A Glaucoma Case Study"

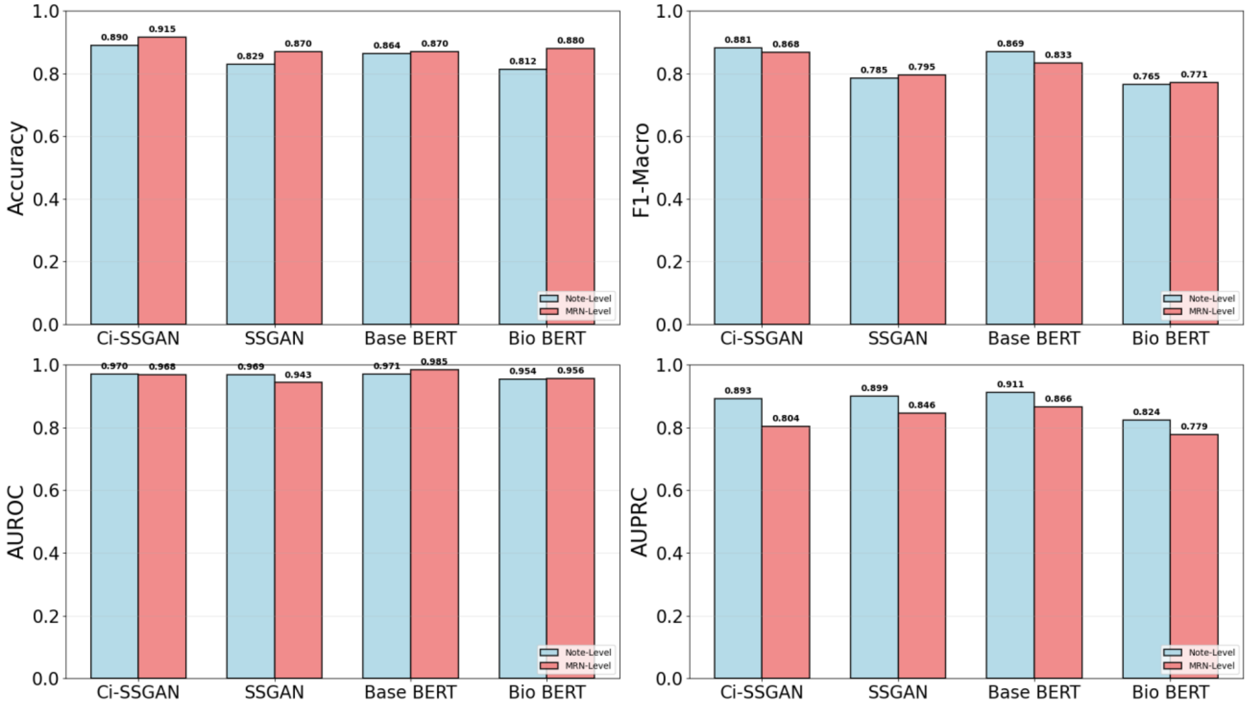


Supplementary Figure 1. Performance comparison of all trained models at 25% of training data. Classification metrics (Accuracy, F1-Macro, AUROC, AUPRC) for Ci-SSGAN versus baseline models under limited data conditions, evaluated at Note-Level (blue) and MRN-Level (red). MRN= Medical Record Number.

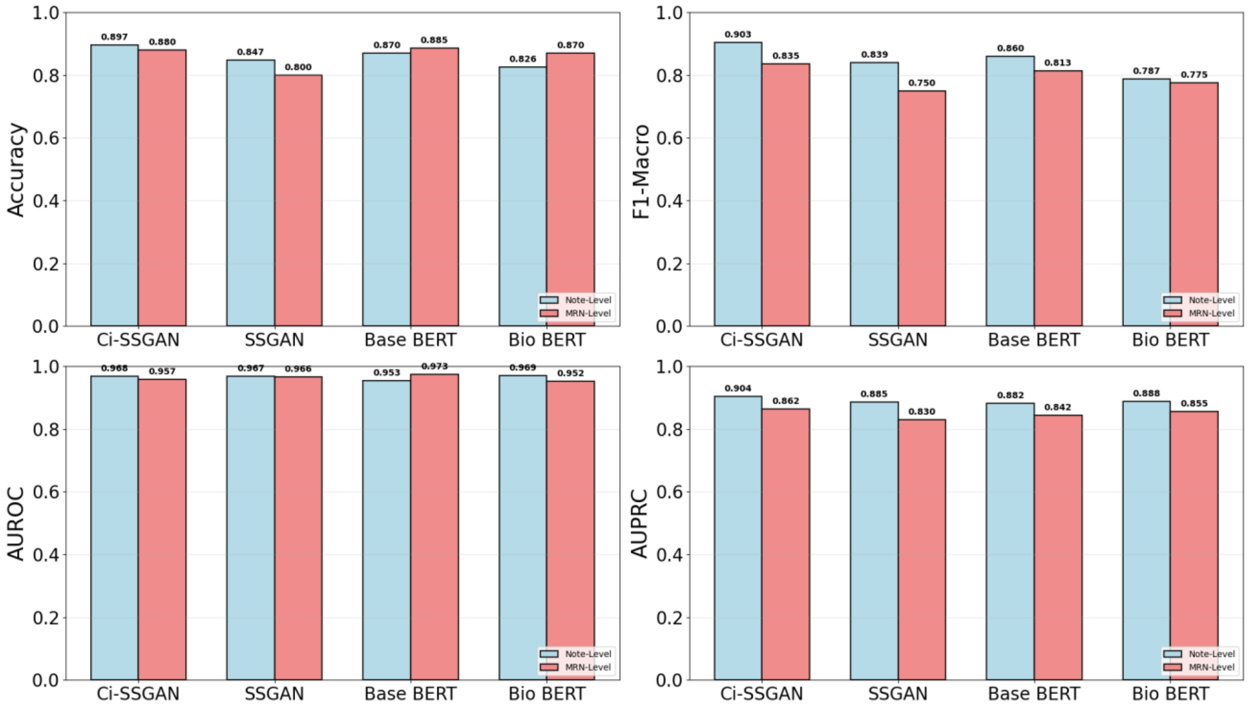


Supplementary Figure 2. Performance comparison of all trained models at 100% of training data. Classification metrics (Accuracy, F1-Macro, AUROC, AUPRC) for Ci-SSGAN versus baseline models under limited data conditions, evaluated at Note-Level (blue) and MRN-Level (red). MRN= Medical Record Number.

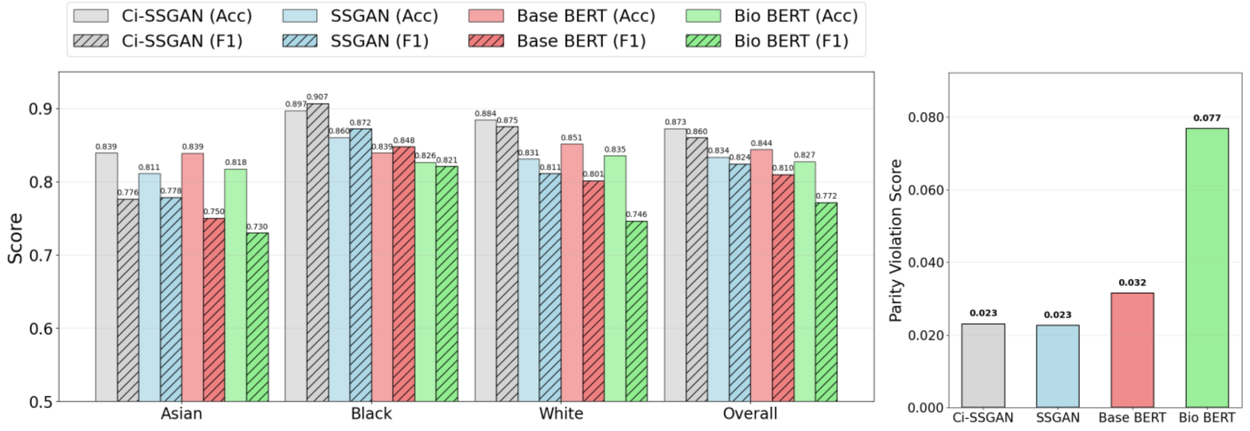


(a)

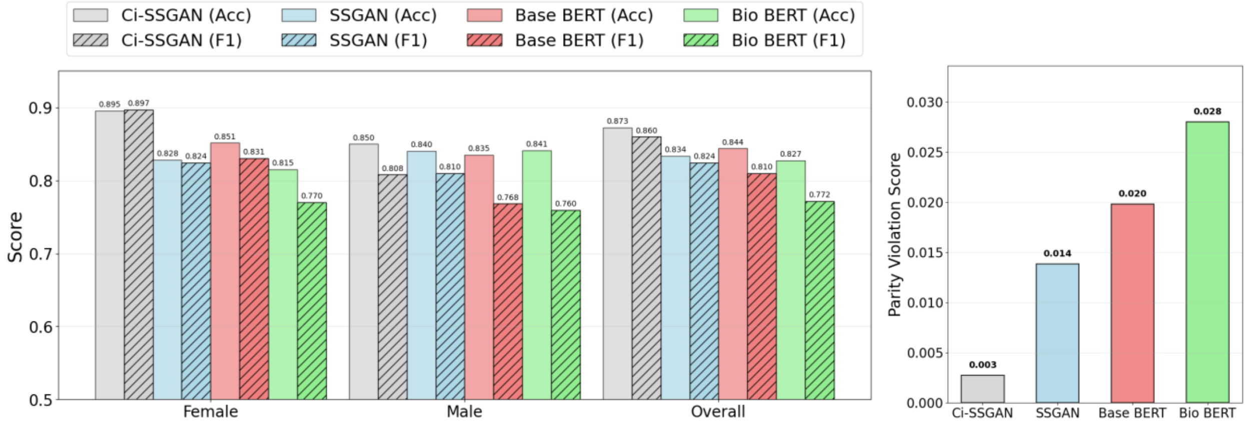

(b)

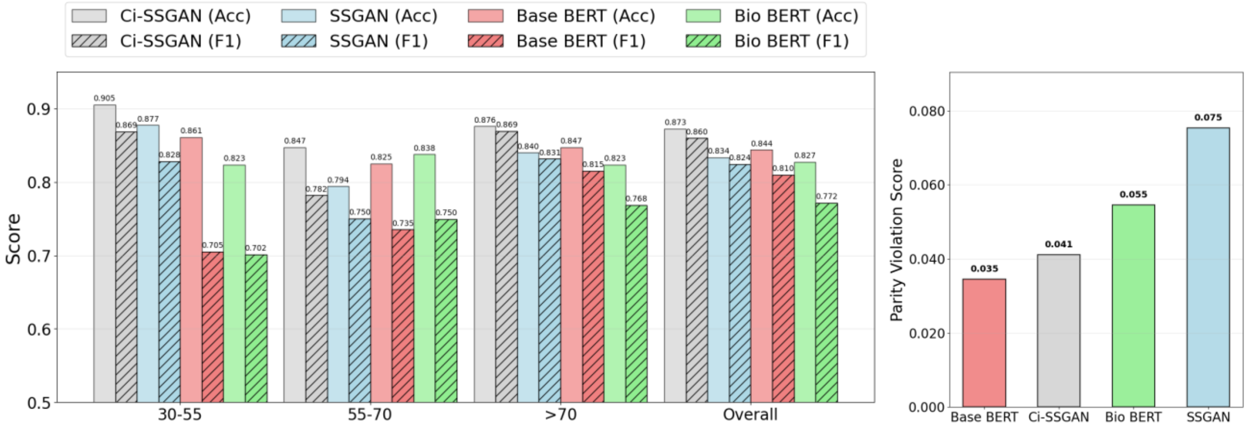


(c)

Supplementary Figure 3. Performance comparison of Ci-SSGAN, SSGAN, Base BERT, and Bio BERT models across **a** Racial groups, **b** Gender groups, and **c** Age groups using 100% of the labeled data. Left panels show accuracy (solid bars) and F1-score (hatched bars) for each subgroup and overall performance. Right panels present corresponding parity violation scores, indicating fairness across demographic subgroups. Ci-SSGAN consistently achieves higher accuracy and F1-scores with lower parity violations compared to other models. The results are presented on five CV folds. Acc= Accuracy, F1= F1- macro.

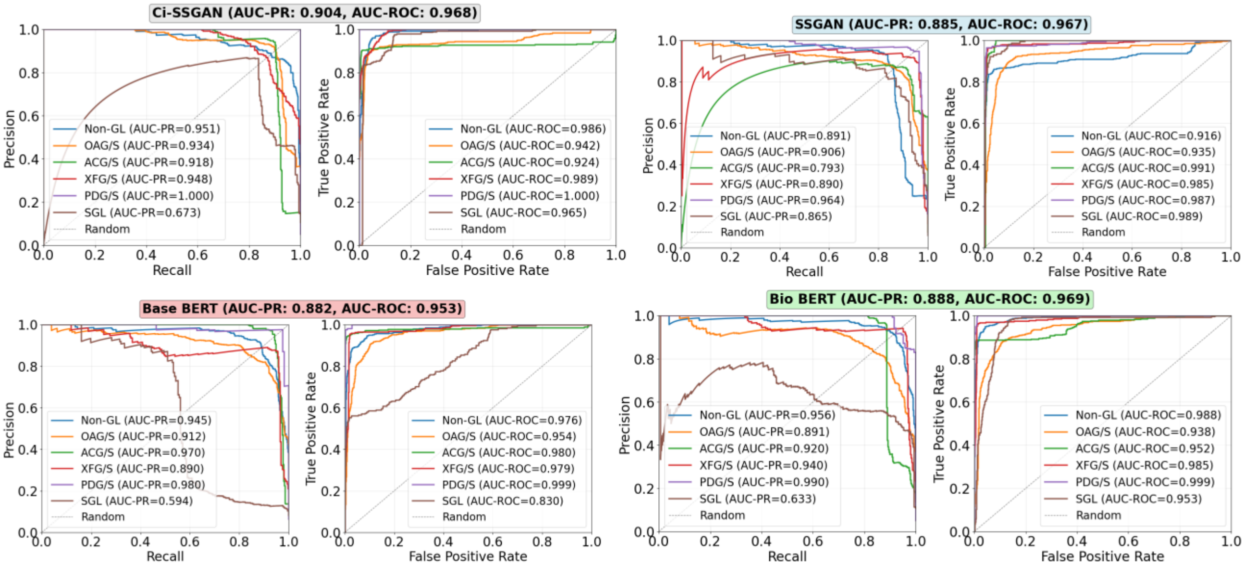


Supplementary Figure 4. Class-wise performance for all trained models. The highest AUC-PR was achieved for Ci-SSGAN with 0.893. The most severely underrepresented class receiving 5× boost factor during training on all four trained models. The results are presented on five CV folds. The dashed line is showing the random classifier. All models were trained using 100% of the labeled data.

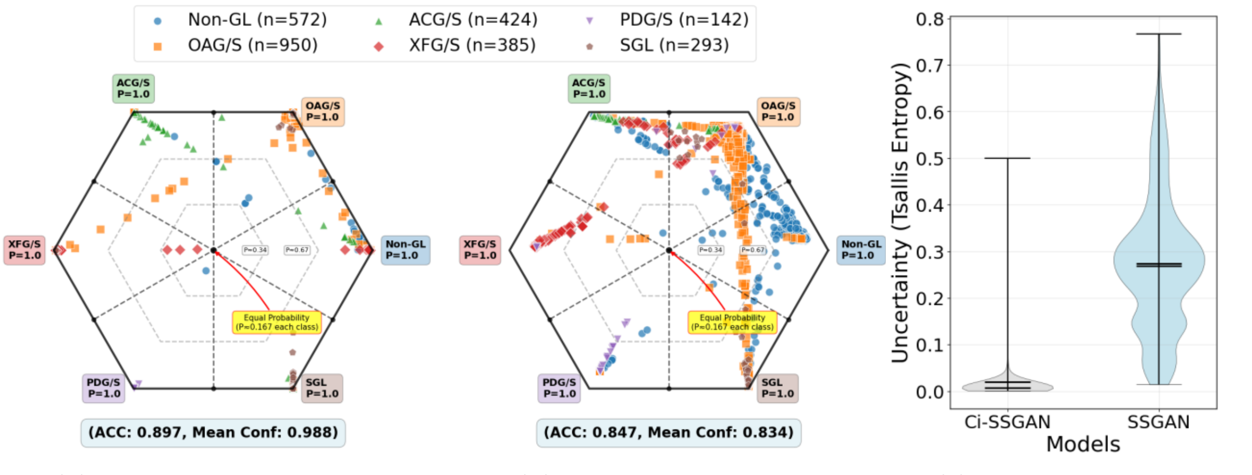


(a) (b) (c)

Supplementary Figure 5. Class probability distributions and predictive uncertainty analysis. **a** Ci-SSGAN radial probability map on the best cv fold. Each vertex represents a class at *P*=1, with concentric rings denoting probability levels from the center at uniform probability (*P*=1/6=0.167) to the outer ring at *P*=1. Point colors correspond to the ground-truth class, and dashed black lines indicate decision boundaries. **b** SSGAN radial probability map using the same visualization scheme. **c** Violin plots of predictive uncertainty (Tsallis entropy). Ci-SSGAN produces lower-entropy, more confident predictions, while SSGAN shows higher-entropy, less certain outputs. High entropy values indicate greater predictive uncertainty, and low values indicate more confident predictions. In the violon plots, the horizontal line indicates the median and the error bars represent the minimum and maximum values observed for each group.

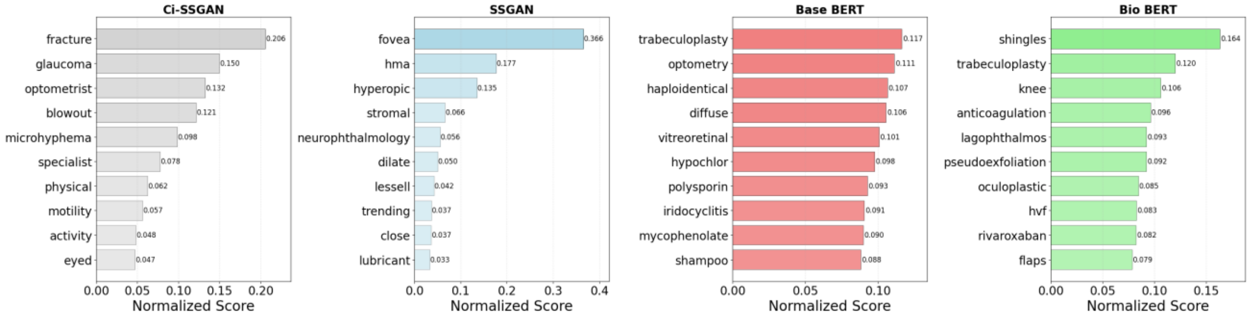


(a)

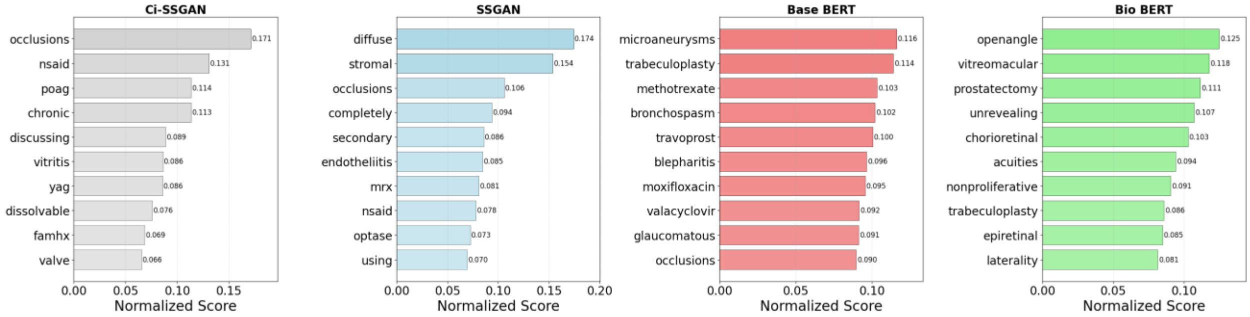


(b)

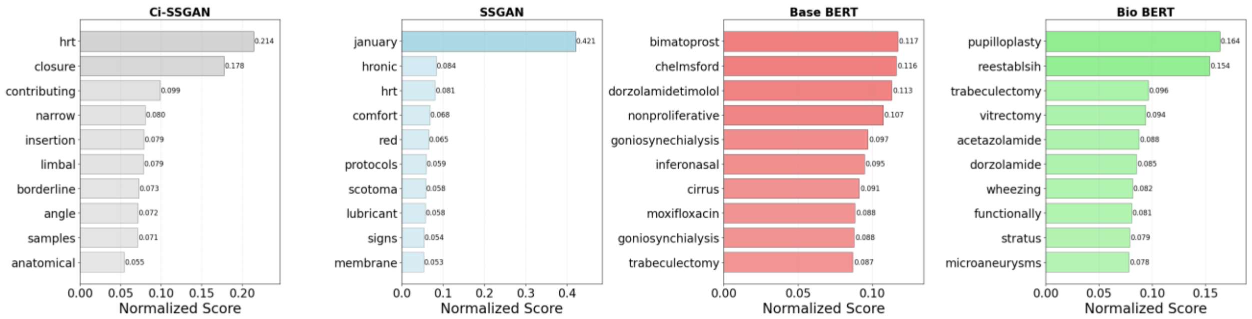


(c)

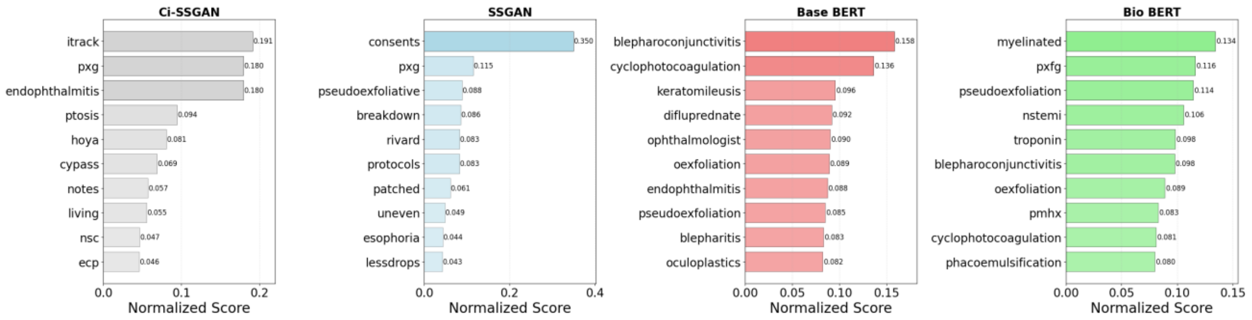


(d)

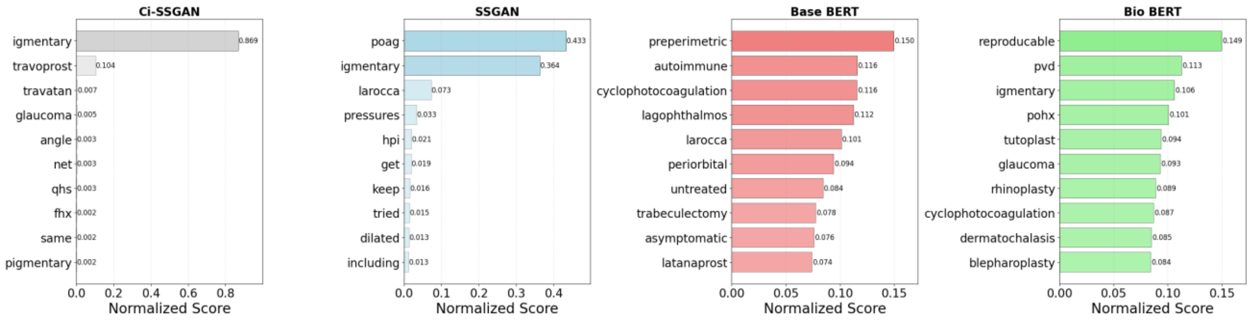


(e)

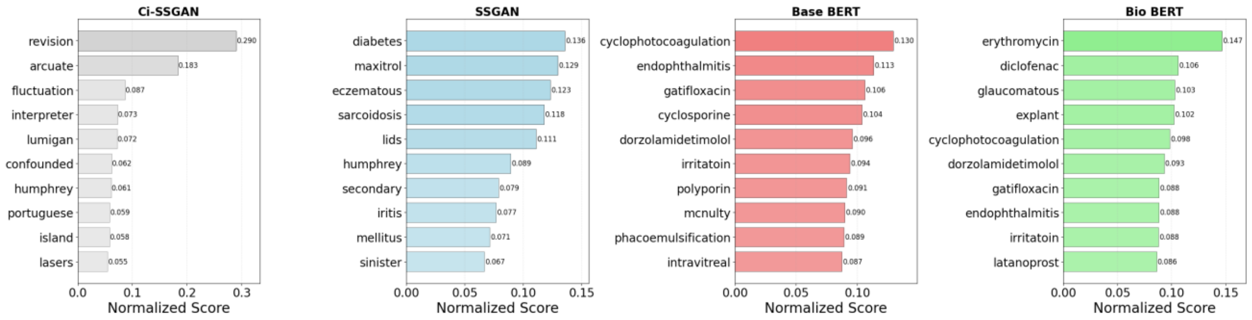


(f)

Supplementary Figure 6. Per class gradient-weighted token attribution analysis across four trained models for glaucoma classification averaged across 5-fold cross-validation. **a** Non-GL, **b** OAG/S, **c** ACG/S, **d** XFG/S, **e** PDG/S, **f** SGL. To have a fair comparison, scores are normalized to sum to 1.0 for each model.

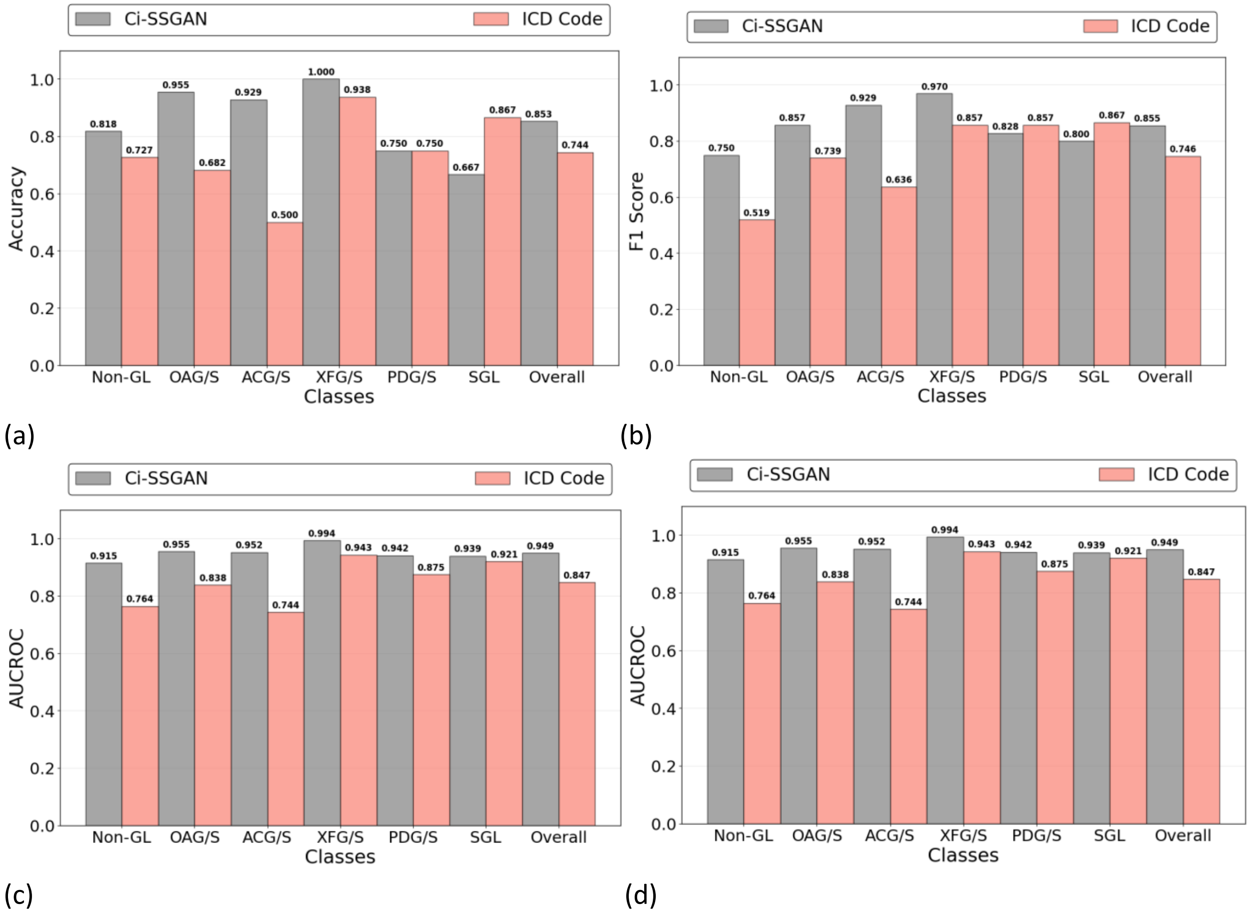


Supplementary Figure 7. Comparison of Ci-SSGAN and ICD code–based labeling across glaucoma subtypes and non-glaucoma cases in terms of **a** Accuracy, **b** F1 Score, **c** AUC-ROC, and **d** AUC-PR. Each bar represents performance for a specific class, with the “Overall” category summarizing all classes. Ci-SSGAN uses both labeled and unlabeled data with clinical context, whereas ICD code labels rely solely on diagnosis codes from medical records. Improvements were most pronounced in challenging subtypes such as primary angle-closure glaucoma (ACG/S), open-angle glaucoma (OAG/S), and Non-GL cases.

Supplementary Table 1. Performance comparison of the Ci-SSGAN at 25% and 100% of the labeled dataset. The values are calculated based on the test data set.

| Metric | Difference (100−25)% | Relative Change (%) |
|---|---|---|
| Accuracy | 0.036 | 4.60 |
| F1 score | 0.039 | 5.10 |
| AUROC | 0.007 | 0.78 |
| AUCPR | 0.026 | 3.17 |

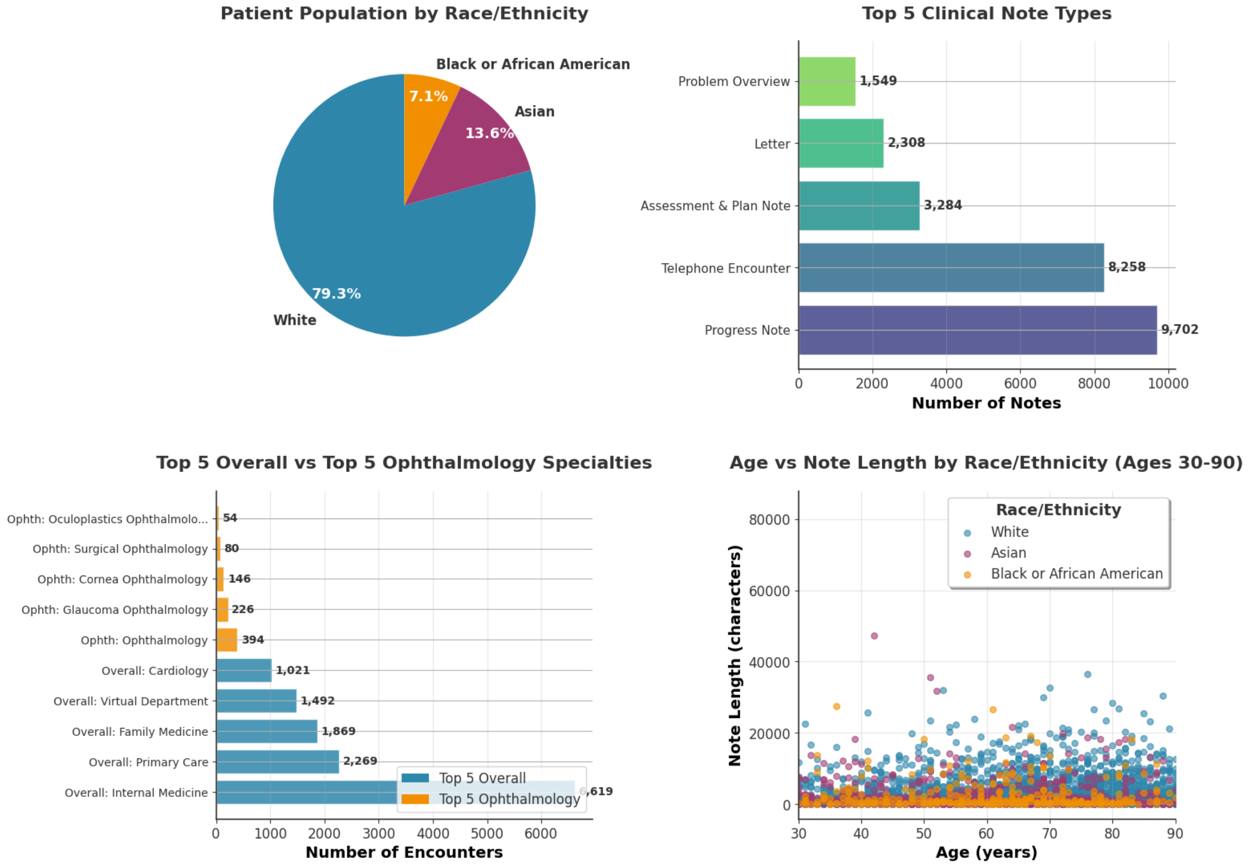


Supplementary Figure 8. Data distribution of the MEE clinical dataset. Top left: Patient population distribution by race/ethnicity showing predominance of White patients (79.3%) among ~1.67M patients. Top right: Top 5 clinical note types by volume, with Progress Notes being the most common (9,702 notes). Bottom left: Comparison of encounter volumes between top 5 overall departments and top 5 ophthalmology specialties, highlighting the specialization in ophthalmology-related encounters. Bottom right: Distribution of note length (in characters) versus patient age (30-90 years) stratified by race/ethnicity, demonstrating consistent documentation patterns across demographic groups with note lengths typically ranging from 0-80000 characters. we have identified 27 distinct note types from 5658 different departments including 224 department specialties with 22.3% of total notes from Internal Medicine. Ophthalmology departments including Ophthalmology, Glaucoma Ophthalmology, Neuro-Ophthalmology, Oculoplastic Ophthalmology, Cornea Ophthalmology, Surgical Ophthalmology, Ophthalmology Imaging, Trauma Ophthalmology, Retinal Degeneration Ophthalmology.

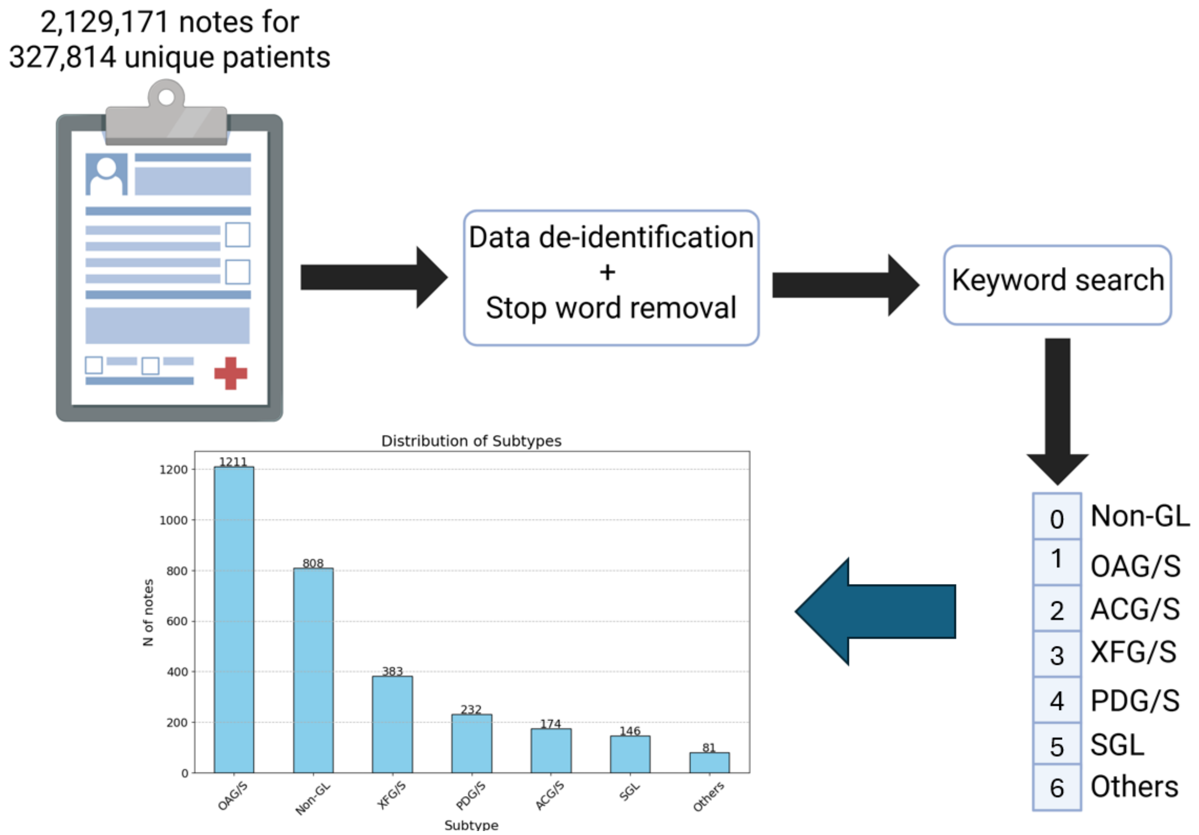


Supplementary Figure 9. Ground truth generation pipeline. The notes were reviewed and graded by 6 independent experts. The labels Non-GL, OAG/S, ACG/S, PDG/S, XFG/S, SGL, and Others are non-glaucomatous open angle glaucoma or suspect, angle closure glaucoma or suspect, pigmentary dispersion glaucoma or syndrome, exfoliation glaucoma or syndrome, secondary glaucoma, and other glaucoma, respectively.

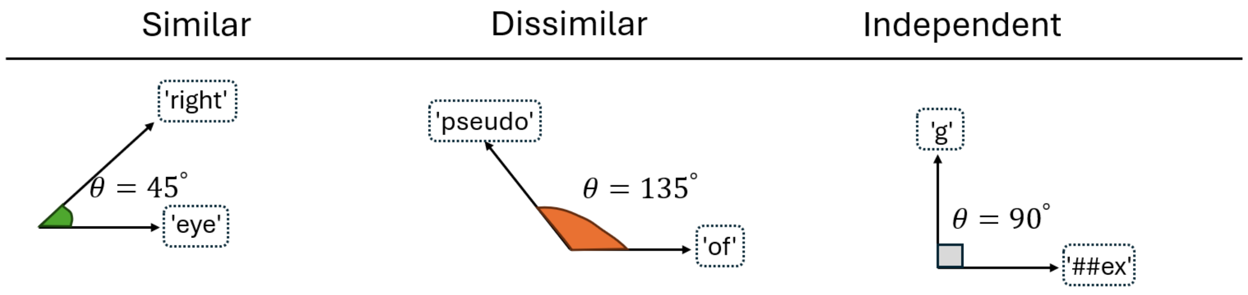


Supplementary Figure 10. Illustration of similar, dissimilar, and independent token pairs used in the generator's diversity loss. The angles (θ) represent cosine similarity between token embeddings, with θ ≈ 45° indicating high similarity, θ ≈ 135° indicating dissimilarity, and θ ≈ 90° indicating orthogonal/independent relationships. Example shown for input text 'Pseudoexfoliation glaucoma of right eye' with tokens: ['pseudo', '##ex', '##folia', '##tion', 'g', '##lau', '##com', '##a', 'of', 'right', 'eye']. This diversity loss ensures the generator maintains meaningful semantic relationships between tokens while preventing mode collapse.

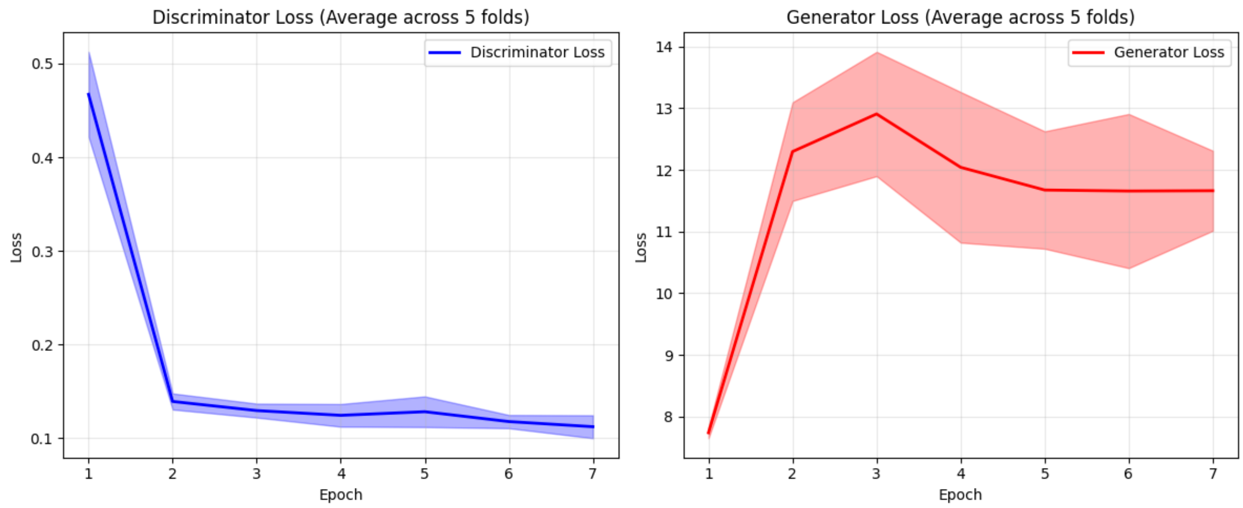


(a) (b)

Supplementary Figure 11. Learning curves for: **a** Generator, and **b** Discriminator. The learning curves demonstrate that the Ci-SSGAN training is stable throughout, with neither the generator nor discriminator losses exhibiting signs of collapse or divergence—hallmarks of successful GAN convergence. The discriminator loss gradually decreases while the generator loss stabilizes over epochs, indicating a healthy adversarial balance. Compared to the baseline BERT model, the SGAN shows improved regularization behavior, as evidenced by smoother loss curves and reduced overfitting. This suggests that the Ci-SSGAN architecture not only maintains training equilibrium but also better generalizes across diverse clinical notes, offering a more robust framework for semi-supervised learning in healthcare NLP tasks.

Supplementary Table 2. The list of loss functions used in the discriminator and generator. $\alpha$ represents class weights, $\gamma = 2$ is the focusing parameter, and $p$ is the predicted probability for the true class. The real and fake losses use binary cross-entropy. The generator loss ($L_G$) incorporates adversarial ($L_{adv}$), feature matching ($L_{feature\_match}$), clinical consistency ($L_{cl\_cons}$), and diversity ($L_{div}$) components. $f_D(\cdot)$ denotes the discriminator feature extraction layers, $z \in \mathbb{R}^{100}$ is the random noise, $d \in \mathbb{R}^3$ represents demographic features, and $x \in \mathbb{R}^{768}$ are unlabeled clinical text embeddings. The feature matching [1,2] loss encourages the generator to match the statistical moments of intermediate feature representations learned by the discriminator. In contrast, the clinical consistency loss operates directly on the generator's output to preserve clinical semantic properties, while diversity loss prevents mode collapse by encouraging dissimilarity between generated samples. diversity loss is also known as cosine similarity loss = $\sum_{ij} \frac{G_i * G_j}{||G_i||_2 * ||G_j||_2}$ which measures similarity between generated embeddings $i$ and $j$, $B$ is the batch size, and $G_i$ is the $i-$th fake text embeddings.

| Loss Component | Definition | Purpose |
|---|---|---|
| **Discriminator loss** | $L_D = L_{supervised} + L_{real} + L_{fake}$ | Combines supervised classification with adversarial training |
| **Supervised loss** | $L_{supervised} = -\alpha(1-p)^{\gamma} \log(p)$ | Addresses class imbalance, focusing on hard-to-classify examples |
| **Generator loss** | $L_G = (L_{adv} + L_{feature\ match} + L_{cl\ cons}) - L_{div}$ | Multi-component loss integrating adversarial, feature matching, clinical consistency, and diversity terms |
| **Adversarial loss** | $L_{adv} = -\log(\sigma(D_{binary}(G(z,d))))$ | Encourages generator to fool discriminator |

| | | |
|---|---|---|
| **Feature matching loss** | $L_{feature\_match} = \|\|E[f_D(x_{real})] - E[f_D(G(z,d,x))]\|\|_2^2$ | Aligns generated features with real data to stabilize training |
| **Clinical consistency loss** | $L_{cl\_cons} = \|\|E[x_{real}] - E[G(z,d,x)]\|\|_2^2$ | reserves medical semantics in generated embeddings |
| **Diversity loss** | $L_{div} = \left(\frac{1}{B(B-1)}\right)\sum_{ij}\frac{G_i * G_j}{\|\|G_i\|\|_2 * \|\|G_j\|\|_2}$ | Encourages generated samples to be distinct, preventing mode collapse |

Supplementary Table 3. Ci-SSGAN complete architecture. The components including the text encoder (12-layer BioClinical BERT), the enhanced generator with clinical context input (871D), and the dual-head discriminator. Parameter counts are shown for each component, with a total of ~112.7M trainable parameters when using full BERT.

| Component | Layer | Input Dim | Output Dim | Parameters |
|---|---|---|---|---|
| **Text Encoder** | Token Embeddings | 28,996 vocab | 768D | 22,268,928 |
| | Position Embeddings | 512 positions | 768D | 393,216 |
| | Token Type Embeddings | 2 types | 768D | 1,536 |
| | **Per Transformer Layers** | | | |
| | Self-Attention | 768 | 768 | 2,362,368 |
| | Feed-Forward | 768 | 768 | 4,722,432 |
| | Layer Norms | 768 | 768 | 3,072 |
| | **× 12 layers=** | | | **85,054,464** |
| | Pooler | 768 | 768 | 590,592 |
| | Attention Pool Layer 1 | 768 | 768 | 590,592 |
| | Attention Pool Layer 2 | 768 | 1 | 769 |
| | **Total Text Encoder=** | | | **109,900,097** |
| **Generator** | FCN 1 | 871 | 1024 | 893,952 |
| | BatchNorm1d | 1024 | 1024 | 2,048 |
| | FCN 2 | 1024 | 768 | 787,200 |
| | BatchNorm1d | 768 | 768 | 1,536 |
| | FCN 3 | 768 | 768 | 590,592 |
| | **Total Generator=** | | | **2,275,328** |
| **Discriminator** | FCN 1 | 771 | 512 | 395,264 |
| | FCN 2 | 512 | 256 | 131,328 |
| | Classifier Head | 256 | 6 | 1,542 |
| | Source Head | 256 | 1 | 257 |
| | **Total Discriminator=** | | | **528,391** |
| **Total Ci-SSGAN** | | | | **112,703,816** |

Supplementary

Supplementary Table 4. Hyperparameters and model configurations used for model training. LR denotes learning rate. Early stopping indicates the patience parameter (number of epochs without improvement before stopping). The noise dimension (100D) refers to the random noise input to the generator, and token size (512) represents the maximum sequence length for text embeddings.

| **Hyperparameters** | **Value** |
|---|---|
| Epochs | 250 |
| Early stopping | 11 |
| Batch size | 16 |
| Generator LR | $2 \times 10^{-4}$ |
| Discriminator LR | $1 \times 10^{-4}$ |
| Text Encoder LR | $3 \times 10^{-5}$ |
| Noise Dimension | 100D |
| Token size | 512 |
